# Risk Factors Associated with Longitudinal Trajectories of Mania Symptoms in Youth

**DOI:** 10.1101/2025.09.03.25334998

**Authors:** Jasmin Dönicke, Rebecca Cooper, Adriane M Soehner, Maria Jalbrzikowski

**Affiliations:** Department of Clinical Psychology and Psychotherapy, Georg Elias Müller Institute of Psychology, Georg August University of Göttingen, Germany; Department of Psychiatry and Behavioral Sciences, Boston Children’s Hospital, Boston, MA, United States; Department of Psychiatry, Harvard Medical School, Boston, MA, United States; Department of Psychiatry, University of Pittsburgh, Pittsburgh, PA, United States

**Author notes:** Corresponding author: Maria Jalbrzikowski, 300 Longwood Ave, Boston MA 02115;, Phone: 617/919-5294. indicates shared first author (Jasmin Dönicke, Rebecca Cooper).

## Abstract

Objective. Mania symptoms in youth predict poor long-term mental health outcomes, yet their developmental trajectories and associated risk factors remain unclear.

Methods. Leveraging data from the Adolescent Brain Cognitive Development Study (N=10,474; 9–10 years at baseline; 48% female, 65% white), we used latent growth mixture models to identify trajectories of mania symptoms across two years in early adolescence. We used multinomial logistic regressions to examine associations between trajectories and risk factors across mental and physical health, cognition, and family/environmental domains.

Results. We identified three trajectories: Low (58%), Moderate (32%), and High/Variable Mania Symptoms (10%). All mental health symptoms (depression, attention deficit hyperactivity disorder, conduct and oppositional defiant disorders and anxiety), two physical health factors (sleep disturbances, irritable bowel syndrome symptoms), one cognitive factor (verbal learning impairment), and three family/environmental factors (trauma, parent- and youth-reported family conflict) significantly differentiated between all three trajectories, reflecting incremental increases in risk factor severity with increasing mania symptoms. Other physical, cognitive, family and environmental factors were also associated with more severe mania symptom trajectories.

Conclusions. More severe mania symptom trajectories in early adolescence are associated with multiple mental, physical, cognitive, family and environmental risk factors, underscoring the need for comprehensive risk prediction approaches in youth. Keywords: Mania Trajectories, Adolescents, Risk Factors, Longitudinal Study, Epidemiology

## Introduction

Symptoms of mania, characterized by increased energy, distractibility, pressured speech, and irritability (Kowatch et al., 2005; Ryles et al., 2017; Van Meter, Burke, Kowatch, et al., 2016), are endorsed by up to 28% of youth and observed across psychiatric disorders (Merikangas et al., 2012; Stringaris et al., 2010). Symptoms of mania are distinguishing features of bipolar disorder, but also occur in youth with depression, anxiety, psychosis, attention-deficit hyperactivity disorder (ADHD) and disruptive behavior disorders (Findling et al., 2010; Geller, Zimerman, et al., 2004; Kim & Miklowitz, 2002). Regardless of diagnostic status, mania symptoms are associated with considerable psychosocial, cognitive, and functional impairment, and increased risk of suicidality (Bechdolf et al., 2014; Lewinsohn et al., 2000; Päären et al., 2013; Stringaris et al., 2010, 2011).

Up to 60% of adults with bipolar disorder report experiencing their first mania symptoms in childhood or adolescence (Chengappa et al., 2003; Lish et al., 1994). However, while mania symptoms may represent prodromal indicators of bipolar disorder, only a subset of youth (2-28%) endorsing these symptoms later receive a diagnosis (Findling et al., 2013; Lewinsohn et al., 2000; Päären et al., 2013; Ratheesh et al., 2023). Longitudinal studies reveal that youth who experience persistent and/or severe mania symptoms are up to six times more likely to later develop bipolar disorder than those who never experience such symptoms, indicating persistent and/or elevated mania symptoms may be prognostic indicators of bipolar risk (Findling et al., 2013; Frazier et al., 2011). Characterizing longitudinal trajectories of mania will aid in early identification and intervention of youth with persistent mania symptoms, ultimately improving long-term outcomes.

Our understanding of mania symptom trajectories is derived primarily from clinical, help-seeking, and familial high-risk samples. The Longitudinal Assessment of Mania Symptoms (LAMS) study assessed over 600 youth engaged in outpatient psychiatric care with elevated mania symptoms over a period of two years, identifying four distinct trajectories characterized by increasing, unstable, rapidly decreasing or gradually decreasing symptoms. Youth with increasing and unstable trajectories were more likely to develop a diagnosis of bipolar disorder than youth with decreasing symptoms (Findling et al., 2013). The Course and Outcomes of Bipolar Youth (COBY) study assessed over 350 youth with bipolar spectrum disorder diagnoses and identified four trajectories based on the proportion of time spent euthymic (i.e., not experiencing a mood episode): predominantly euthymic, moderately euthymic, ill with improving course and predominantly ill. Youth with a predominantly ill course experienced the most severe mania symptoms, the longest manic episodes, and the greatest severity of psychiatric comorbidity (Birmaher et al., 2014). In a parallel line of work, several studies have assessed mania symptoms in youth with a family history of bipolar disorder. These studies demonstrate that mania symptoms are common and frequently precede bipolar disorder onset in familial high-risk youth (Axelson et al., 2015; Duffy et al., 2019).

However, while these studies provide valuable insights into mania symptom trajectories among help-seeking, clinical or familial high-risk populations, these samples often included youth that met criteria for psychiatric disorders at baseline, limiting the generalizability to community samples. Developmental trajectories of mania symptoms in representative, population-based samples are poorly understood - yet such data is essential for advancing early detection and intervention efforts.

Mania symptoms and bipolar disorders are associated with a range of psychiatric, physical, cognitive and environmental risk factors (Supplemental Table 1). These include mental health symptoms related to depression, anxiety, ADHD and behavioral disorders (Brancati et al., 2021; Buckley et al., 2023; Jackson et al., 2003); physical concerns including sleep disturbances, irritable bowel syndrome, asthma, obesity, and a history of birth and obstetric complications (Melo et al., 2016; Shintani et al., 2023; Tseng et al., 2016; Wu et al., 2016; Zhao et al., 2016) and impairments in verbal learning and memory (Kurtz & Gerraty, 2009). A family history of bipolar disorder, as well as adverse early experiences such as trauma and maltreatment and are also established risk factors for mania and bipolar disorders (Agnew-Blais & Danese, 2016; Mortensen et al., 2003). However, much of this evidence is derived from cross-sectional or retrospective studies conducted in adults (Tseng et al., 2016; Wu et al., 2016; Zhao et al., 2016), or in clinical samples (Brancati et al., 2021; Jackson et al., 2003). Of studies conducted in youth, many have been cross-sectional (e.g. Tseng et al., 2015); providing limited insight into their role in predicting longitudinal symptom trajectories.

Moreover, the relative strength of associations between risk factors and symptom trajectories, and their utility in identifying youth with severe and persistent symptoms, is currently unknown.

In this study, we identified longitudinal trajectories of mania symptoms in the diverse and representative Adolescent Brain and Cognitive Development study (Volkow et al., 2018). First, we used latent trajectory analyses to characterize distinct patterns of mania symptom progression over time by considering both the severity and course of mania symptoms across repeated assessments. We identified 20 established risk factors for mania and bipolar disorder and examined associations between trajectory group membership and the established risk factors. Lastly, we assessed the overall variance explained by including all risk factors as predictors of trajectory membership.

## Methods

### Participants

We analyzed data from the Adolescent Brain Cognitive Development (ABCD) study, a longitudinal cohort study examining brain development and child health in the United States (Volkow et al., 2018). The ABCD study recruited over 11,800 youth, aged 9-10 years at baseline, from 21 geographically and demographically diverse research sites across the US. We included data from baseline, 1-year follow-up, and 2-year follow-up (5.1 release). We excluded participants if they had missing data on our primary measure, the Parent General Behavior Inventory, at baseline or at any follow-up assessment (N = 1,234). We also excluded participants who had a diagnosis of bipolar disorder at baseline (N = 136), resulting in a final sample of 10,434 youth (48% female, 52% male). We used the Kiddie Schedule for Affective Disorders and Schizophrenia for DSM-5 (Kaufman et al., 1997; Townsend et al., 2020) to determine bipolar disorder diagnosis. The study’s protocols were approved by institutional review boards at each participating site, with parents or guardians providing informed consent and the youth providing assent.

## Measures

### Mania symptoms

We assessed mania symptoms via the 10 Item Mania Scale, derived from the Parent General Behavior Inventory (PGBI-10M; Youngstrom et al., 2008), a 10-item parent-report scale designed to assess hypomania and mania symptoms in youth (refer to the Supplemental Methods and Supplemental Table 1 for alternative measures).

Parents report on their child’s behaviors in the past year, scoring each item on a scale from 0 (never or hardly ever) to 3 (very often or almost constantly), yielding a total score ranging from 0 to 30, with higher scores reflecting greater symptom severity. Scores less than 5 indicate low risk for bipolar disorder; scores between 5 and 9.9 indicate low to moderate risk, 10 to 14.9 indicate moderate risk, 15 to 17.9 indicate high risk, and scores 18 and above indicate very high risk for bipolar disorder (Youngstrom et al., 2008). We chose the PGBI-10M after conducting a review of all potential mania questionnaires used in the ABCD. See Supplemental Methods for additional details.

Because the peak age of onset for bipolar disorder is 19.5 years (Solmi et al., 2022), and the peak age of onset for a manic episode is 21-25 years (Kennedy, Everitt, et al., 2005), we did not examine categorical diagnoses of manic episodes or bipolar spectrum disorders as either predictor or outcome variables.

### Identification of Risk Factors

We conducted a literature review to identify empirically-validated risk factors for mania and/or bipolar disorder. We searched PubMed and PsycINFO for studies that analyzed factors associated with onset of mania symptoms or diagnosis of bipolar disorder and were published in the last two decades (2004-2024). We selected risk factors that (1) were not identical to symptoms of subsequent manic episodes, (2) were a statistically significant risk factor for mania or bipolar disorder in at least two independent studies, (3) had a conceptually matching variable within the ABCD, and (4) had sufficient data collected for this variable at the baseline assessment. We identified 20 risk factors associated with mental health (i.e., depressive, anxiety, attention deficit hyperactivity disorder [ADHD], oppositional defiant disorder, conduct disorder symptoms), physical health (i.e., sleep disturbances, irritable bowel syndrome [IBS] symptoms, asthma, body mass index [BMI], pregnancy and obstetric complications, premature birth, low birth weight, and Cesarean delivery), cognition (verbal learning impairment), and family/environmental factors (family conflict, childhood trauma, maltreatment, and first-degree relatives with mania). Full details of the literature review can be found in Supplemental Table 2; selected risk factors are detailed in Supplemental Table 3.

#### Mental Health Risk Factors

We used the parent-reported DSM-5-oriented scales from the Child Behavior Checklist (Achenbach, 1991) to assess depression, anxiety, ADHD, oppositional defiant, and conduct disorder symptoms over the past six months.

#### Physical Health Risk Factors

Sleep disturbances were assessed with the 26-item Sleep Disturbance Scale for Children (SDSC; Bruni et al., 1996). Parents reported on their child’s sleep behaviors over the past six months. We calculated a total score from these responses (possible range 26-130), with higher scores indicating greater sleep disturbance. In post-hoc analyses, we also calculated six subscale scores that address specific sleep disorder domains: sleep initiation and maintenance disorders, breathing disorders, arousal disorders, sleep-wake transition disorders, excessive somnolence, and hyperhidrosis. We evaluated IBS-related symptoms using three parent-reported responses to questions about nausea, stomach aches, and vomiting from the Child Behavior Checklist (Achenbach, 1991; Supplemental Table 3). Similar to previous work (Kerr et al., 2021), we summed these items into a total score (range 0-6), with higher scores indicating more frequent or severe gastric symptoms. We assessed asthma with a parent-reported question from the ABCD Medical History Questionnaire: "Has she/he ever been to a doctor for asthma?". We calculated BMI using height and weight. We excluded outliers that were >5 standard deviations above the mean BMI. Parents completed the ABCD Developmental History Questionnaire, which assesses pregnancy and birth complications, such as premature birth, low birth weight (indicated by birth weight <2.5kg; Shintani et al., 2023), or birth via Cesarean delivery (Barch et al., 2018). Specific variables used are detailed in Supplemental Table 3.

#### Cognitive Risk Factors

We measured verbal learning using the Rey Auditory Verbal Learning Test (RAVLT; Schmidt, 1996). Participants were asked to recall fifteen words across five trials. For each trial, the number of correctly recalled words (range 0-15) was recorded. We summed these scores across trials to create a total correct score (range 0-75), which we then reverse-scored. Higher scores indicate greater verbal learning impairment.

#### Family and Environment Risk Factors

The ABCD study measured family conflict using the Family Conflict subscale from the Family Environment Scale (Moos & Moos, 1976). Youth and parents separately rated nine items as true or false, generating sum scores ranging from 0-9, with higher scores indicating greater conflict. Childhood trauma was assessed using the 17-item parent-rated Kiddie Schedule for Affective Disorders and Schizophrenia - Post-Traumatic Stress Disorder (KSADS-PTSD; Kaufman et al., 1997) scale. The total score, summed from all yes/no items and ranging from 0-17, indicates greater trauma exposure (Thomas et al., 2024). Childhood maltreatment was assessed using a 13-item composite measure based on previous work by Warrier et al. (2021), yielding a total score ranging from 0 to 13. The items were drawn from two sources: (1) eight items from the parent-reported KSADS-PTSD (Kaufman et al., 1997) module, capturing various forms of abuse and trauma exposure, and (2) five items from the youth-reported Children’s Report of Parental Behavior Inventory (Schaefer, 1965), re-coded to reflect emotional neglect (see Supplemental Table 3; Perkins et al., 2024). Only responses indicating “Not like him/her” were classified as reflecting potential emotional neglect due to low parental warmth, contributing to the overall maltreatment score. We used responses from the ABCD Family History Questionnaire to determine the presence of mania in first-degree relatives (Barch et al., 2018).

#### Covariates

Parents/caregivers provided information on sex assigned at birth (male/female), Hispanic ethnicity and race from 15 available categories. Consistent with previous work (Fung et al., 2023; Hiraoka et al., 2023), we consolidated the ABCD race variable into six categories (American Indian/Alaskan, Asian, Black/African American, Pacific Islander, White, Other/Multiple). We assessed socioeconomic status using the income-to-needs ratio (Gonzalez et al., 2020).

## Statistical Analyses

We used growth mixture models (Mplus version 8.11; Muthén & Muthén, 1998-2025) to characterize mania symptom trajectories. Due to the high positive skewness and excess zeroes in the data (Supplemental Figure 1), we ran both zero-inflated Poisson and negative binomial models to identify the optimal modeling approach (Table 2). Following recommendations (Mara & Carle, 2021; Nylund et al., 2007; Ram & Grimm, 2009), we estimated models with one to six classes for each model type (Poisson and negative binomial) and determined the best fitting model using the following qualitative and quantitative criteria (Jung & Wickrama, 2008): lower Bayesian information criterion (BIC) values, class-specific posterior probabilities high for one class (ideally >0.85) and near zero for other classes, entropy values closer to one, class proportions that encompassed >5% of the sample, model parsimony, and previous literature. We visualized trajectories identified in each model to ensure classes showed distinct patterns.

**Table 1.** Descriptive statistics for overall sample and mania trajectories.

| **Characteristic** | **Overall** | **High/Variable**<br>**Symptoms** | **Moderate**<br>**Symptoms** | **Low Symptoms** |
| --- | --- | --- | --- | --- |
| N (%); Mean (SD) | N = 10,474 | N = 1,111 | N = 3,316 | N = 6,047 |
| Sex assigned at birth |  |  |  |  |
| Male | 5,482 (52%) | 634 (57%) | 1,745 (53%) | 3,103 (51%) |
| Female | 4,992 (48%) | 477 (43%) | 1,571 (47%) | 2,944 (49%) |
| Age (baseline, months) | 118.98 (7.51) | 118.71 (7.53) | 118.96 (7.51) | 119.04 (7.50) |
| Race |  |  |  |  |
| American Indian/Alaskan | 49 (0.5%) | 7 (0.6%) | 22 (0.7%) | 20 (0.3%) |
| Asian | 217 (2.1%) | 10 (0.9%) | 55 (1.7%) | 152 (2.5%) |
| Black/African American | 1,507 (14%) | 260 (23%) | 496 (15%) | 751 (12%) |
| Pacific Islander | 14 (0.1%) | 0 (0%) | 5 (0.2%) | 9 (0.1%) |
| White | 6,834 (65%) | 598 (54%) | 2,112 (64%) | 4,124 (68%) |
| Other/Multiple | 1,833 (18%) | 232 (21%) | 619 (19%) | 982 (16%) |
| Hispanic ethnicity | 2,019 (20%) | 260 (24%) | 701 (21%) | 1,058 (18%) |
| Income to Needs Ratio | 403.69 (282.91) | 267.22 (252.19) | 377.13 (273.21) | 441.41 (284.22) |
| Mania symptom score |  |  |  |  |
| N (%); Mean (SD) | N = 10,474 | N = 1,111 | N = 3,316 | N = 6,047 |
| Baseline | 1.22 (2.65) | 6.44 (4.88) | 1.54 (1.58) | 0.08 (0.27) |
| 1-year follow-up | 1.30 (2.78) | 7.08 (4.91) | 1.57 (1.5) | 0.09 (0.29) |
| 2-year follow-up | 1,09 (2.52) | 5.93 (4.79) | 1.34 (1.57) | 0.06 (0.24) |

**Table 2.**
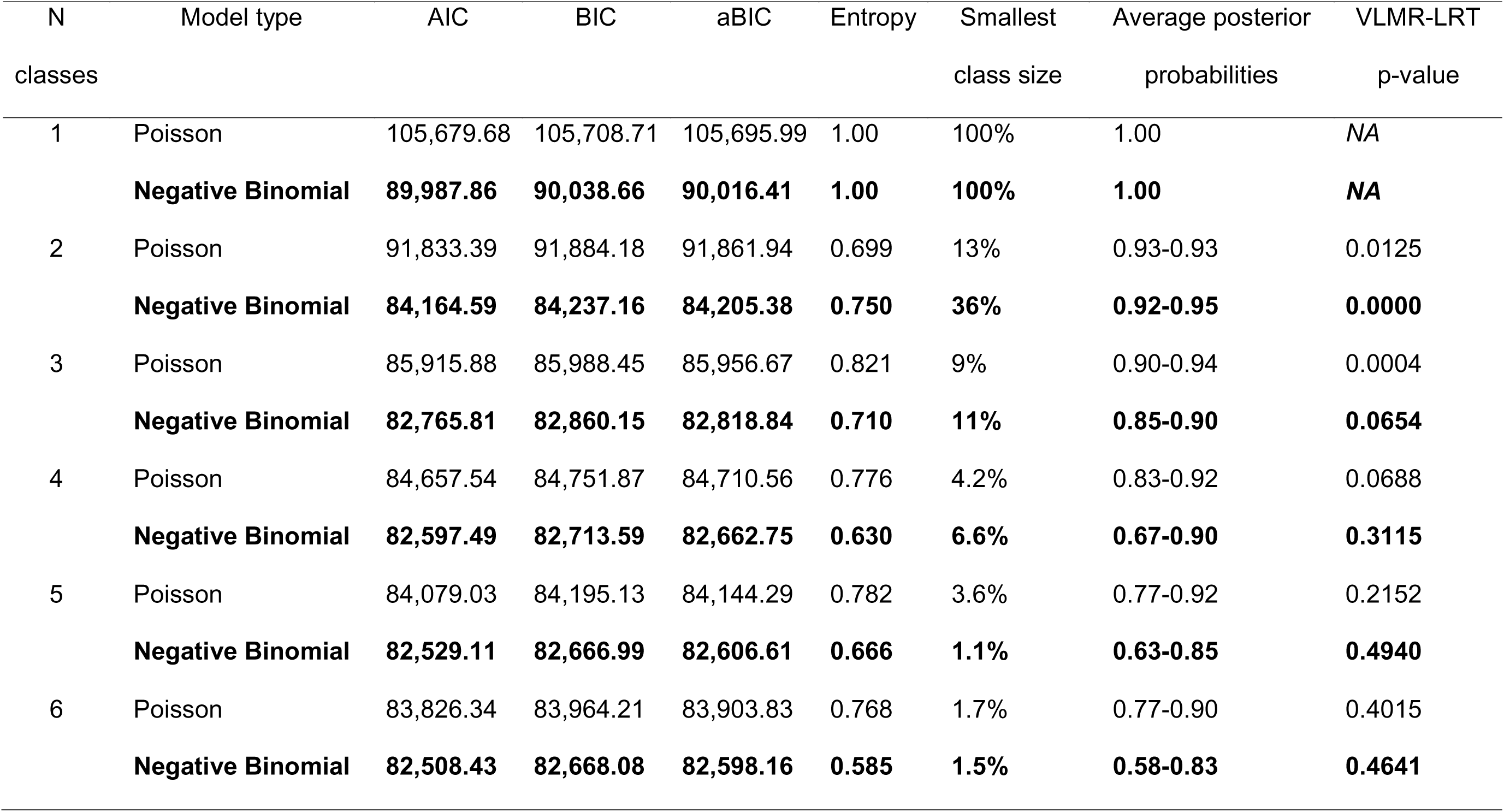
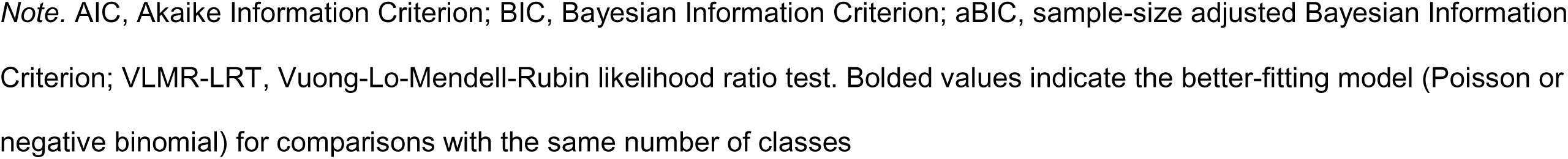
Model fit statistics for linear zero-Inflated Poisson and negative binomial growth mixture models of mania symptoms.

| N | Model type | AIC | BIC | aBIC | Entropy | Smallest | Average posterior | VLMR-LRT |
| --- | --- | --- | --- | --- | --- | --- | --- | --- |
| classes |  |  |  |  |  | class size | probabilities | p-value |
| 1 | Poisson | 105,679.68 | 105,708.71 | 105,695.99 | 1.00 | 100% | 1.00 | NA |
|  | **Negative Binomial** | **89,987.86** | **90,038.66** | **90,016.41** | **1.00** | **100%** | **1.00** | **NA** |
| 2 | Poisson | 91,833.39 | 91,884.18 | 91,861.94 | 0.699 | 13% | 0.93-0.93 | 0.0125 |
|  | **Negative Binomial** | **84,164.59** | **84,237.16** | **84,205.38** | **0.750** | **36%** | **0.92-0.95** | **0.0000** |
| 3 | Poisson | 85,915.88 | 85,988.45 | 85,956.67 | 0.821 | 9% | 0.90-0.94 | 0.0004 |
|  | **Negative Binomial** | **82,765.81** | **82,860.15** | **82,818.84** | **0.710** | **11%** | **0.85-0.90** | **0.0654** |
| 4 | Poisson | 84,657.54 | 84,751.87 | 84,710.56 | 0.776 | 4.2% | 0.83-0.92 | 0.0688 |
|  | **Negative Binomial** | **82,597.49** | **82,713.59** | **82,662.75** | **0.630** | **6.6%** | **0.67-0.90** | **0.3115** |
| 5 | Poisson | 84,079.03 | 84,195.13 | 84,144.29 | 0.782 | 3.6% | 0.77-0.92 | 0.2152 |
|  | **Negative Binomial** | **82,529.11** | **82,666.99** | **82,606.61** | **0.666** | **1.1%** | **0.63-0.85** | **0.4940** |
| 6 | Poisson | 83,826.34 | 83,964.21 | 83,903.83 | 0.768 | 1.7% | 0.77-0.90 | 0.4015 |
|  | **Negative Binomial** | **82,508.43** | **82,668.08** | **82,598.16** | **0.585** | **1.5%** | **0.58-0.83** | **0.4641** |

*Note.* AIC, Akaike Information Criterion; BIC, Bayesian Information Criterion; aBIC, sample-size adjusted Bayesian Information Criterion; VLMR-LRT, Vuong-Lo-Mendell-Rubin likelihood ratio test. Bolded values indicate the better-fitting model (Poisson or negative binomial) for comparisons with the same number of classes

To assess how baseline risk factors predicted trajectory membership, we used a bias-adjusted three-step approach, which accounts for misclassification in class assignment when modeling covariates in mixture models (Asparouhov & Muthén, 2014). This approach mitigates bias in estimating associations with external variables (e.g. baseline predictors), particularly when entropy is low (i.e. <0.8; Asparouhov & Muthén, 2014). This approach involves: (1) estimating the latent class model without covariates, (2) assigning each individual to their most likely latent class, and (3) using multinomial logistic regression to assess the relationship between class assignment and predictor variables while accounting for the misclassification from the second step (Asparouhov & Muthén, 2014). In Step 3, separate models were run for each risk factor variable, adjusting for covariates including sex, race, ethnicity, and socioeconomic status. We applied False Discovery Rate (FDR) correction to account for multiple comparisons across the 20 risk factors (Benjamini & Hochberg, 1995). Results are reported as adjusted Odds Ratios (aORs) with 95% Confidence Intervals (CIs), using the lowest severity mania symptom class as the reference category.

We compared the strength of associations between mental health, physical health, cognitive, family and environmental, and sociodemographic risk factors with mania trajectory membership by including all risk factors in a multivariable logistic regression model predicting trajectory membership (represented by a latent variable). We assigned a model parameter to each group of risk factors, and ran pairwise tests of equality of parameter coefficients using Wald tests of the linear hypothesis *H*_0_: β_1_ - β_2_ = 0, where β_1_ and β_2_ represent the parameter coefficients for each group of risk factors. The Wald test follows a ^2^ distribution with one degree of freedom (Agresti, 2013). Due to missingness on risk factor variables, we restricted this analysis to individuals with complete data across all risk factors (N=8,603).

## Results

Descriptive statistics for the sample are provided in Table 1. Mean mania symptom scores are presented in Table 1 and Supplemental Figure 2.

## Mania symptoms show diverse trajectories over time

We selected a three-class growth mixture model using a negative binomial distribution as the optimal model. Negative binomial models consistently provided a better fit than Poisson models (based on lower BIC; Table 2). BIC values decreased with an increasing number of classes; however, models with five and six classes showed trajectories that were indistinguishable or had low posterior probabilities (see Supplemental Figure 3). In comparing the three- and four-class solutions, we favored the three-class solution due to its higher entropy (three-class 0.71 vs. four class 0.63) and higher posterior probabilities (0.85-0.90 vs. 0.67-0.90). The four-class solution also had a non-significant p-value for the VLMR-LRT (p=0.31), indicating this solution was not a significantly better fit than the three-class model. Additionally, this model included two low severity classes with minimal differences in mania symptom scores, suggesting that they could be combined to achieve a more parsimonious solution. We thus selected the three-class solution as the optimal model (Figure 1). We labeled the first trajectory group "High/Variable Mania Symptoms" (N = 1,111; 10%). There was considerable individual variability within this trajectory group; while some individuals had high symptom scores across all three time points, others experienced severe mania symptoms at only one or two time points (see Supplemental Figure 4). We named the second trajectory group "Moderate Mania Symptoms" (N = 3,316; 32%), which consisted of individuals with moderately elevated mania symptoms that decreased slightly over time (slope term β = -0.09, p = 0.011). We labeled the third trajectory group "Low Mania Symptoms" (N = 6,047; 58%), which included individuals with relatively low symptoms that decreased throughout the study (slope term β = -0.170, p < 0.001).

**Figure 1.**
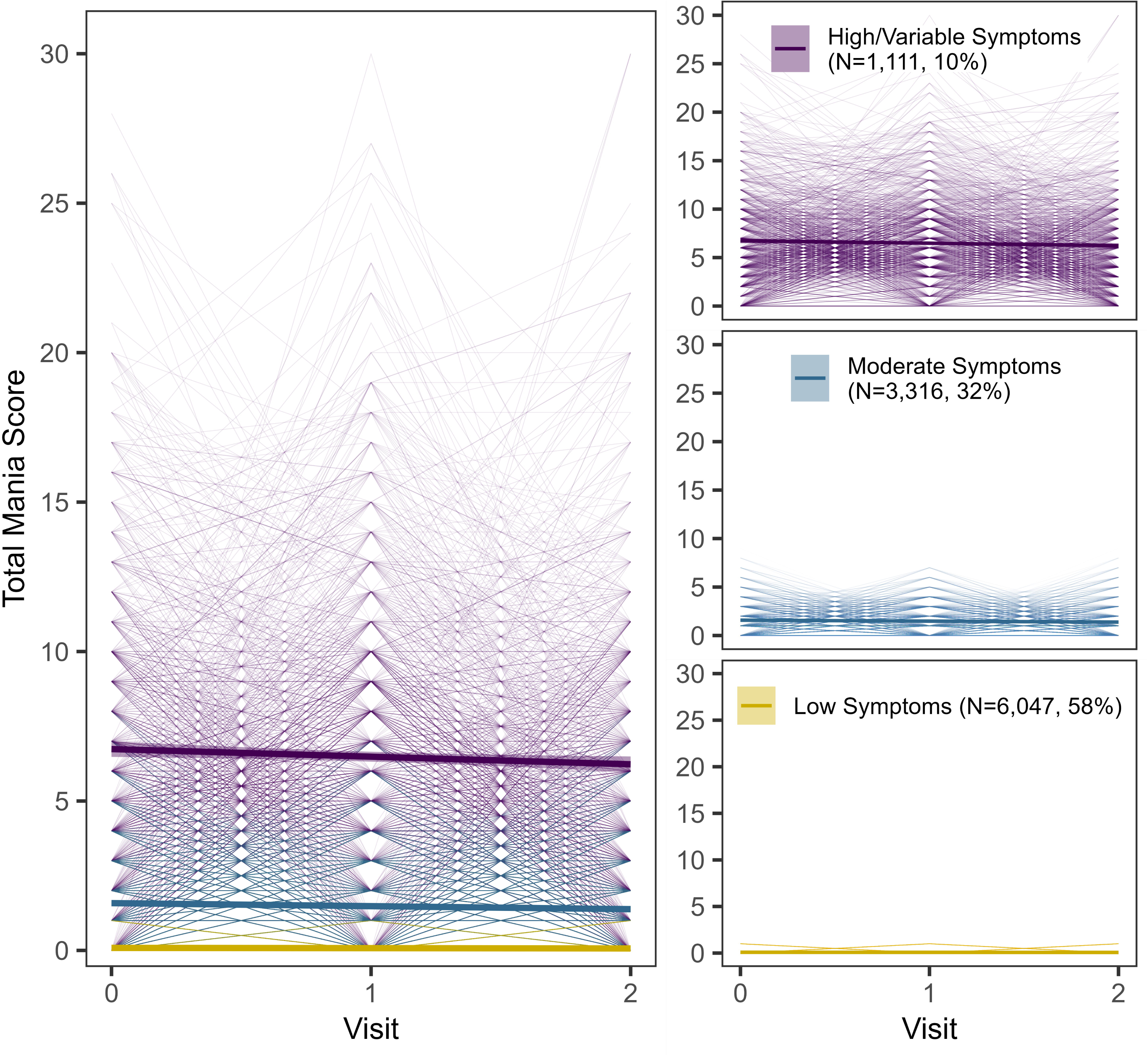
Mania Trajectories with Individual Subject Variability. The left panel presents each mania trajectory. Thick lines represent mean scores for each trajectory; shading represents 95% confidence intervals. Thin lines represent individual scores across study visits. The right panel shows each trajectory group, with bold lines representing mean scores and shading representing 95% confidence intervals. Thin lines represent individual scores across study visits.

## Multiple mental health risk factors for mania and bipolar disorder are associated with mania trajectory group membership

Established risk factors for mania significantly predicted trajectory membership, with greater risk linked to more severe and variable mania symptoms (Figure 2, Table 3 & Supplemental Table 4). For mental health factors, individuals in the High/Variable trajectory experienced greater severity of ADHD, depression, ODD, conduct disorder and anxiety symptoms compared to all other trajectories (aOR range 1.86-10.56). For all mental health symptoms, the Moderate Symptoms trajectory showed greater symptom severity than the Low Symptoms trajectory (aOR range 2.55-4.04). Overall, all mental health symptoms followed a clear stepwise gradient, increasing in severity across the Low Symptoms to the High/Variable Symptoms trajectory.

**Figure 2.**
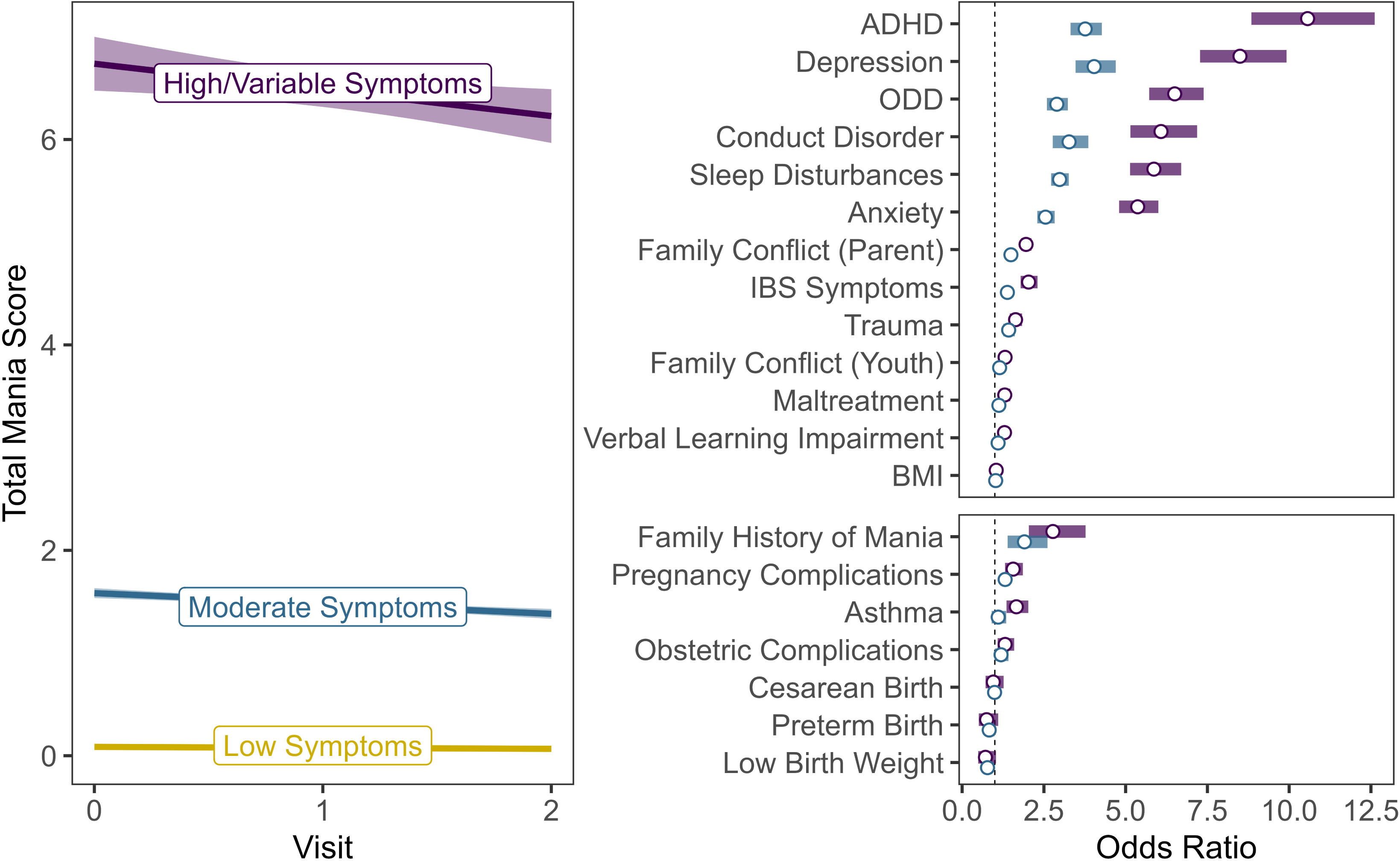
Associations Between Mania Trajectories and Risk Factors. The left panel shows each mania trajectory. Thick lines represent mean score for each trajectory; shading represents 95% confidence intervals. The right panel presents odds ratios (circles) and 95% confidence intervals (bars) for associations between mania trajectories and risk factors, using the Low Symptoms trajectory as the reference group. The top panel shows continuous predictors; the bottom panel shows binary predictors. All models are adjusted for sex, race, ethnicity, and socioeconomic status. Abbreviations: ADHD, attention deficit hyperactivity disorder; BMI, body mass index; IBS, irritable bowel syndrome; ODD, oppositional defiant disorder.

**Table 3.**
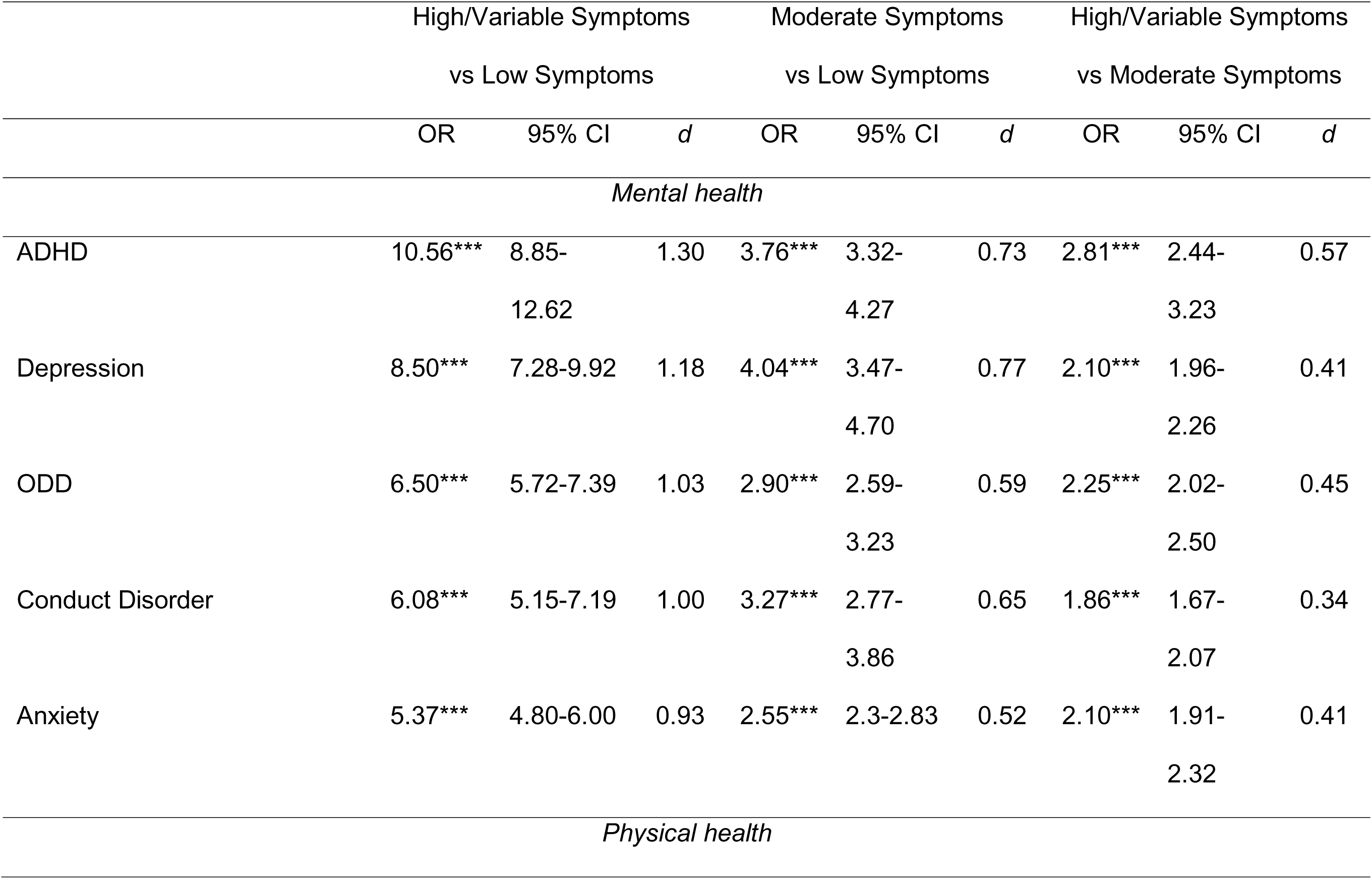
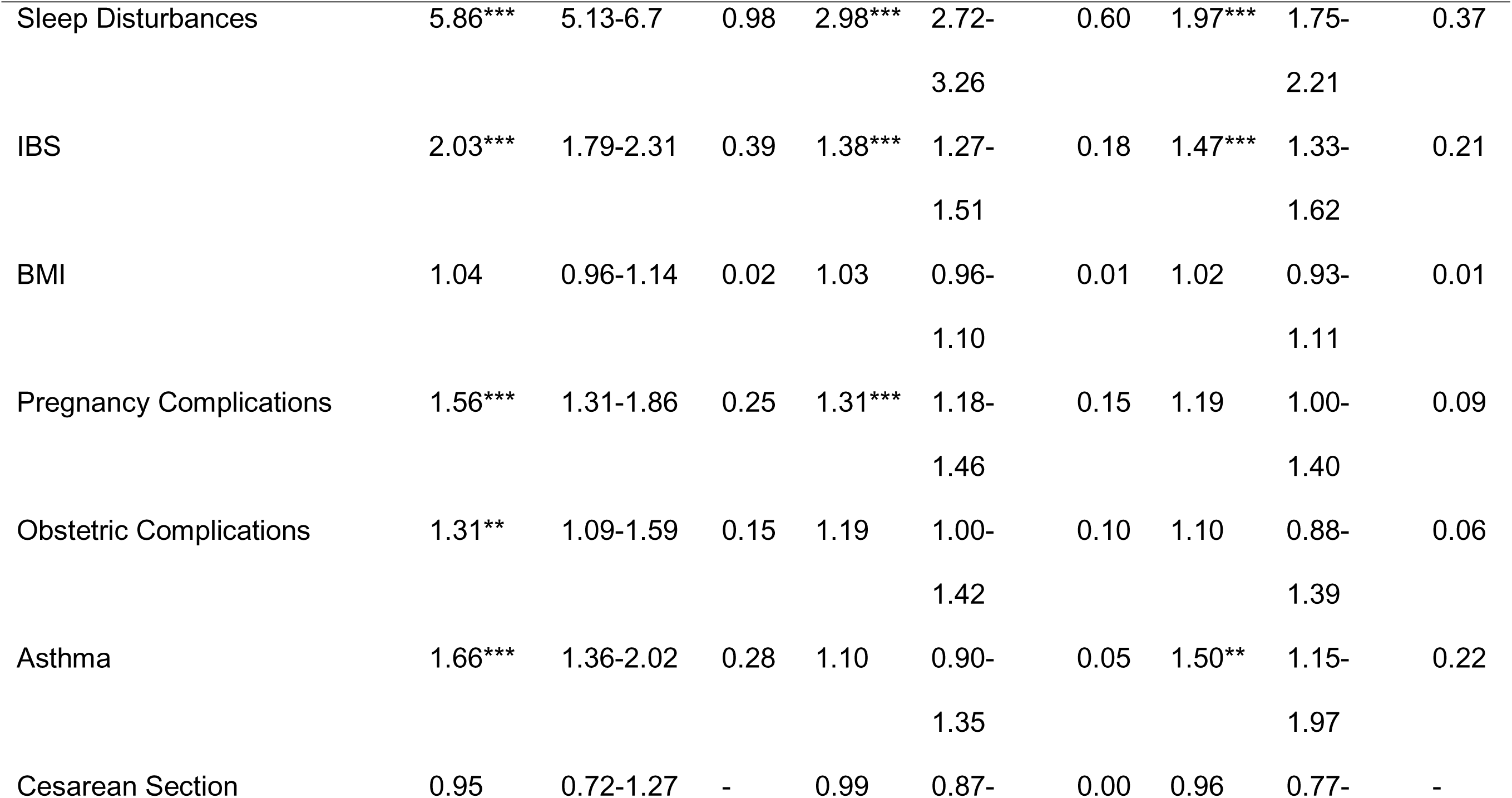
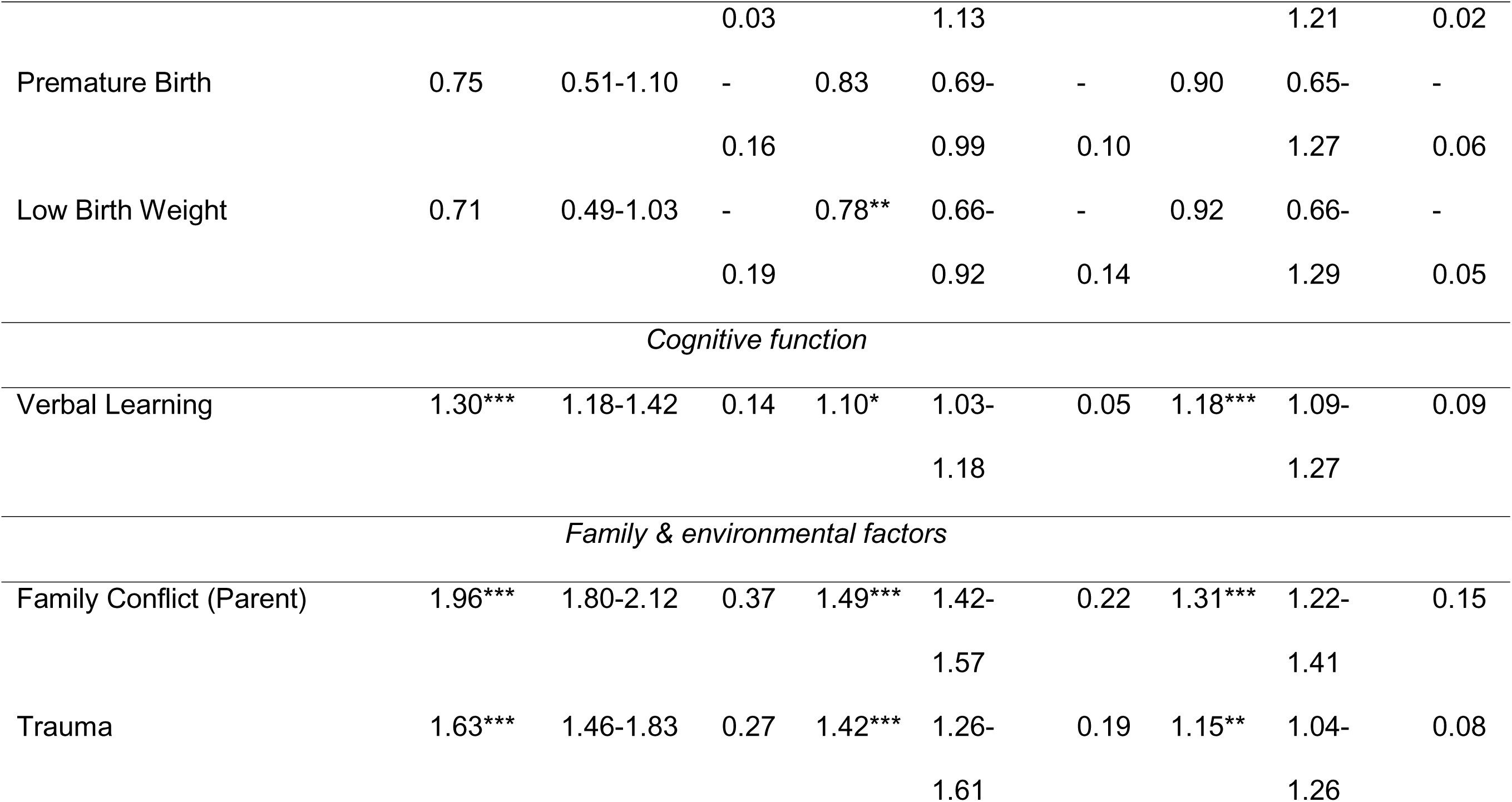
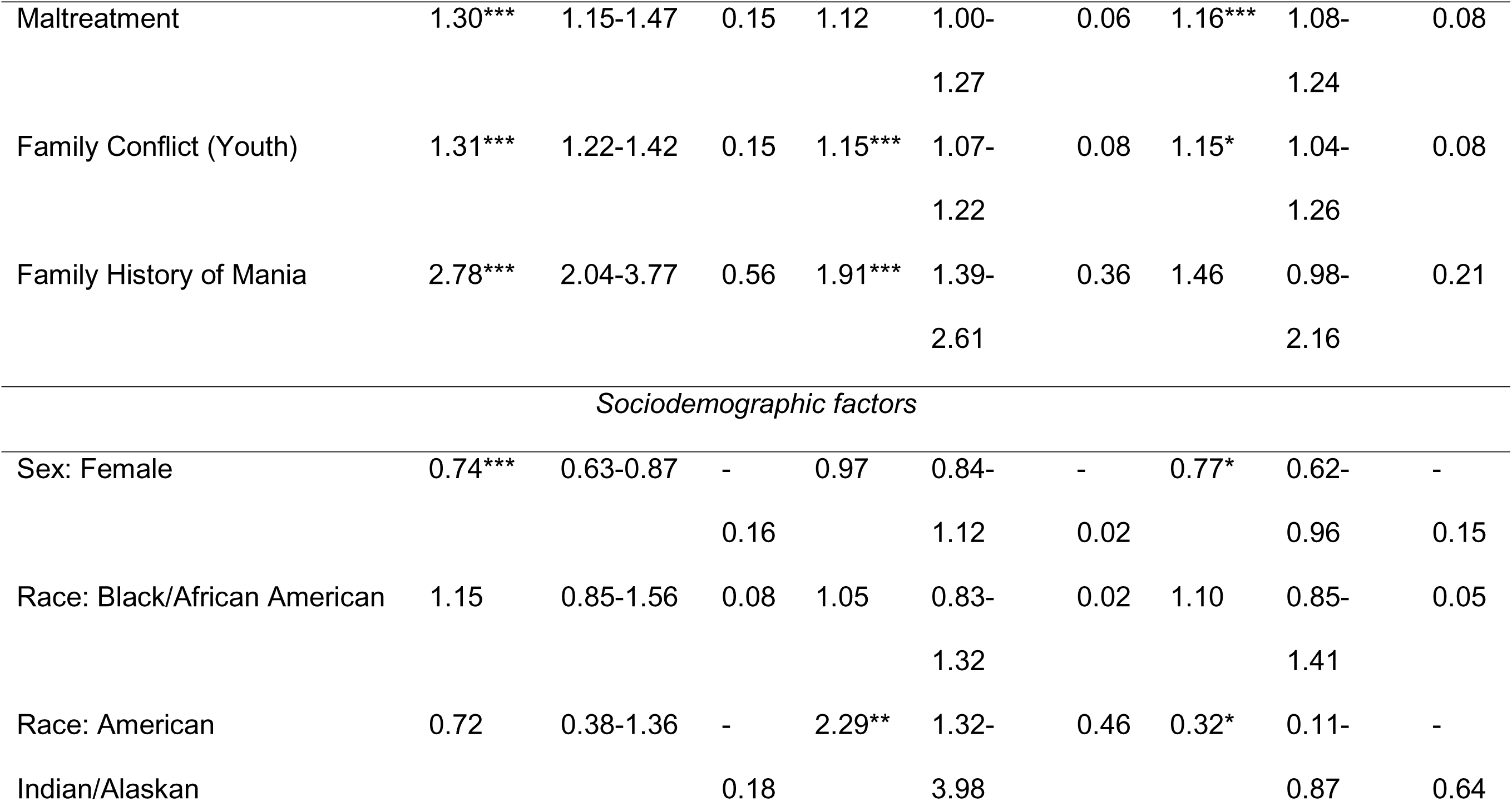
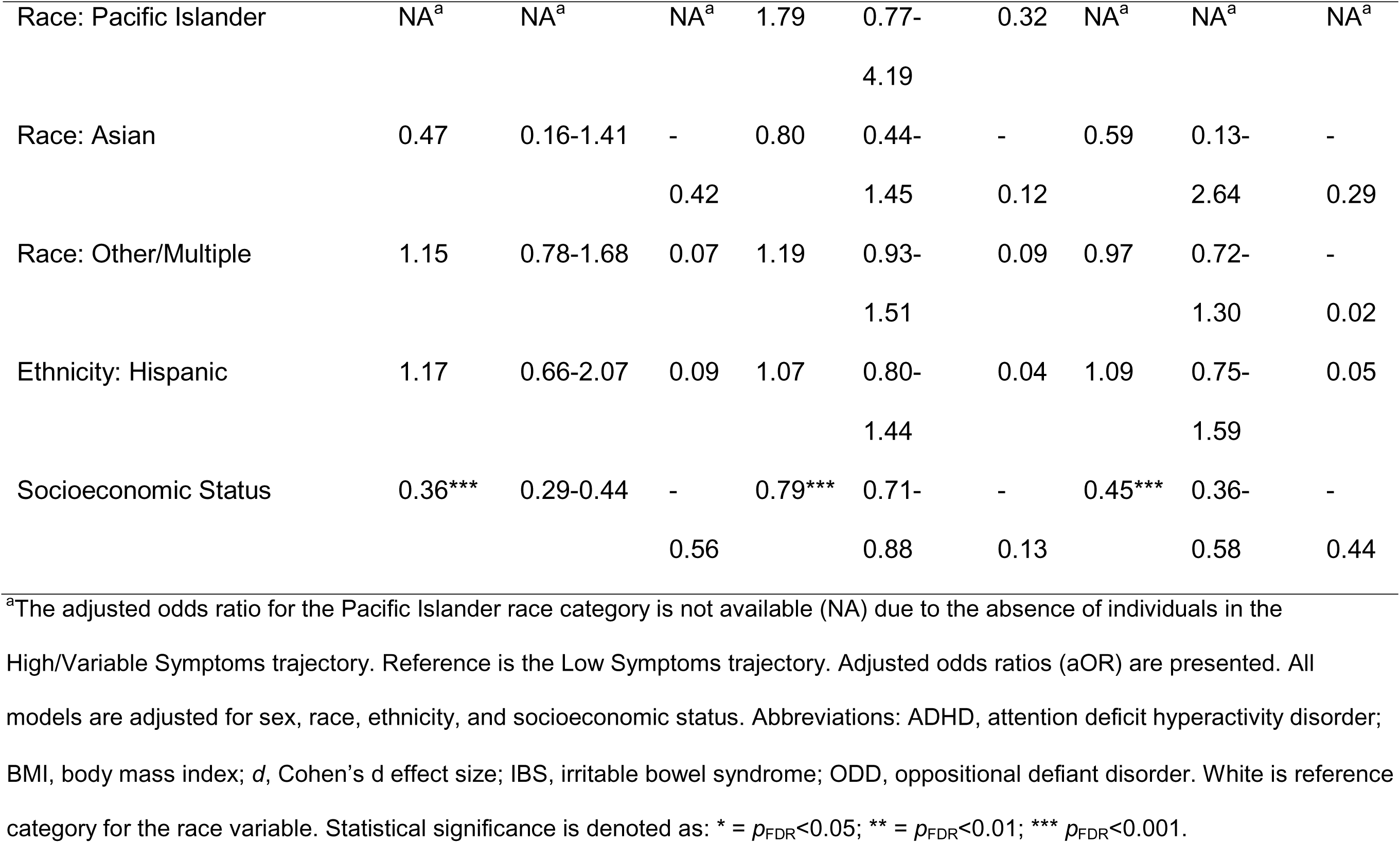
Associations between risk factors and mania trajectories.

For physical health risk factors, sleep disturbances and IBS symptoms also showed a stepwise gradient pattern, with symptoms incrementally increasing in severity across the Low Symptoms to the High/Variable Symptoms trajectories. Individuals in the High/Variable and Moderate Symptoms trajectories showed greater severity of maternal pregnancy complications than the Low Symptoms trajectory (aOR range 1.31-1.56). Individuals in the High/Variable Symptoms trajectory were also more likely to endorse asthma and maternal obstetric complications than those in the Low Symptoms trajectory (aOR range 1.31-1.66). The Moderate Symptoms trajectory showed more severe sleep disturbances, IBS symptoms and maternal pregnancy complications than the Low Symptoms trajectory (aOR range 1.31-2.98). BMI, premature birth, and Cesarean section birth were not associated with trajectory membership.

In terms of cognitive risk factors, verbal learning impairments significantly differentiated between the High/Variable, Moderate and Low Symptom trajectories, with increasing mania symptom severity associated with greater verbal learning impairments (aOR range 1.10-1.30).

Regarding family and environmental risk factors, the High/Variable and Moderate Symptoms trajectories were more likely to endorse trauma, a family history of mania, and parent- and youth-reported family conflict than those in the Low Symptoms trajectory (aOR range 1.15-2.78). The High/Variable Symptoms trajectory also had greater endorsement of maltreatment relative to the Low Symptoms trajectory (aOR 1.30, *p*_FDR_<.001), and greater parent- and youth-reported family conflict, trauma, and maltreatment than the Moderate Symptoms trajectory (aOR range 1.15-1.31). Trauma, parent- and youth-reported family conflict significantly differentiated between the three trajectories, with greater severity of mania symptoms corresponding to greater severity of family and environmental risk factors.

## Mental health and sociodemographic factors were most strongly associated with trajectory group membership

Wald tests showed that coefficients for mental health risk factors were stronger than physical health, family/environmental, and cognitive risk factors in predicting mania trajectory membership (Supplemental Figure 5, Supplemental Table 4). Combined, sociodemographic risk factors were more strongly associated with trajectory membership than cognitive and physical health factors, but were not statistically different from mental health or family/environmental risk factors in predicting trajectory membership. Family and environmental risk factors were comparable to physical health factors in predicting trajectory membership.

## Specific sleep behaviors are strongly associated with mania symptom trajectories

In a post-hoc analysis, we examined associations between the six sleep disorder subscales of the Sleep Disorder Scale for Children and trajectory group membership (Supplemental Table 6, Supplemental Figure 5). All subscales showed a stepwise gradient pattern, with incremental and statistically significant increases in sleep disturbances across the Low Symptoms to the High/Variable Symptoms trajectories.

Numerically, we observed the greatest effect sizes for Disorders of Initiating and Maintaining Sleep, Disorders of Excessive Somnolence, and Sleep-Wake Transition Disorders in differentiating each group.

## Discussion

We identified three unique longitudinal trajectories of mania symptoms in a diverse community sample of over 10,000 U.S. adolescents. Our study is the first to examine trajectories of mania symptoms in relation to established risk factors for mania and bipolar disorder. An array of mental health symptoms (including ADHD, depression, oppositional defiant disorder, conduct disorder and anxiety), physical health factors (sleep disturbances, IBS symptoms, pregnancy and obstetric complications and asthma), cognitive factors (verbal learning impairments) and family- and environment-related risks (having a first-degree relative with mania, exposure to family conflict, trauma and childhood maltreatment) emerged as significant predictors of mania symptom trajectories. Several mental health, physical health, cognitive and family and environmental factors significantly differentiated between all three trajectories, reflecting increased mania symptom severity with incremental increases in ADHD, depression, ODD, conduct disorder, anxiety, sleep disturbances, IBS symptoms, verbal learning impairments, family conflict and trauma. These findings demonstrate the value of leveraging established risk factors for mania and bipolar disorder to identify youth at greatest risk for developing elevated mania symptoms, which has potential to improve risk prediction of severe psychiatric disorders and inform early intervention approaches.

Our analysis identified three distinct mania trajectory classes, comprising two relatively stable/unchanging trajectories and one severe/variable symptom trajectory. These distinct patterns differentiate mania symptoms from many other forms of psychopathology, which show more gradual symptom changes. For example, within the ABCD cohort, internalizing and externalizing symptoms show increasing, persistent, decreasing and low severity trajectories, whereas psychotic-like experiences show severe and persistent, rapidly decreasing, gradually decreasing, and low symptom trajectories (Brieant et al., 2025; Cooper et al., 2025). The presence of a High/Variable trajectory also aligns with previous work that identified severe and unstable mania symptom trajectories across a 24-month period among help-seeking youth (Findling et al., 2013). Both severe and unstable trajectories identified in prior work also experienced more severe depressive symptoms and poorer psychosocial functioning, highlighting that the High/Variable trajectory identified in our work may be most likely to benefit from early identification and intervention.

Mental health risk factors emerged as the strongest predictors of elevated mania symptoms. All mental health symptoms – ADHD, depression, conduct and oppositional defiant disorder and anxiety – showed a stepwise gradient of severity, demonstrating distinct separation of trajectory classes. Consistent with previous work, ADHD and depression emerged as the strongest risk factors, followed by associations with conduct and oppositional defiant disorder (Carlson et al., 2000; Kennedy, Boydell, et al., 2005; Morcillo et al., 2012; Sandstrom et al., 2021). Symptoms of anxiety were also associated with mania trajectory membership, which is consistent with findings that nearly half of individuals with bipolar disorder experience comorbid anxiety (Buckley et al., 2023; Gilman et al., 2012; Johnson et al., 2000). On the other hand, the *pattern* of associations we observed differentiates mania symptom trajectories from other forms of psychopathology. For example, trajectories of psychotic-like experiences in young adolescents are associated with lower symptoms of anxiety, and are not linked to ADHD, depressive or conduct disorder symptoms (Jia et al., 2025), whereas irritability symptom trajectories show strong associations with ADHD, followed by weaker associations with anxiety, depressive and conduct disorder symptoms (Bellaert et al., 2025). Together, these findings suggest that mania symptoms in young adolescents often occur amidst a spectrum of comorbid transdiagnostic psychopathology, and the unique clusters of transdiagnostic symptoms may assist with identifying clinical markers specific to mania. Assessment of comorbid psychiatric symptoms may be particularly valuable in younger adolescents, as comorbid conditions are associated with an earlier onset of bipolar disorder (Carlson et al., 2000; Perlis et al., 2004). Since bipolar disorder often remains undiagnosed until later stages, integrating routine screening for mania symptoms into clinical assessments—particularly for children with ADHD and depressive symptoms—may improve diagnostic accuracy and early treatment planning (Duffy et al., 2016).

Individuals with more physical health factors were more likely to be assigned to the High/Variable and Moderate Symptoms trajectories. Consistent with previous research (Liang & Chikritzhs, 2013; Liu et al., 2015; Melo et al., 2016; Shintani et al., 2023; Tseng et al., 2016; Wu et al., 2016), we found that sleep disturbances and IBS symptoms were associated with more severe mania symptoms, and further differentiated between all three trajectories. In our post-hoc analyses, disorders of initiating and maintaining sleep emerged as the strongest sleep-related factor, followed by disorders of excessive somnolence. Notably, such sleep-disordered symptoms are characteristic of depressive disorder, underscoring the strong interplay between mania and depressive symptoms (Comsa et al., 2022). However, unlike prior studies (Shintani et al., 2023), Cesarean section and premature birth were not significantly associated with mania trajectory group membership. This discrepancy could be due to prior work examining risk for bipolar disorder as distinct from mania symptoms more generally. As only a subset (2-28%) of individuals who experience mania later develop bipolar disorder, our findings suggest pregnancy and obstetric complications may be more relevant to bipolar disorder diagnoses (Findling et al., 2013; Lewinsohn et al., 2000; Päären et al., 2013; Ratheesh et al., 2023). We also did not observe an association between obesity and mania trajectories; thus, like other studies, obesity may be more closely linked to depressive symptoms rather than manic symptoms in bipolar disorder (Goldstein et al., 2011; Kim et al., 2009).

Individuals with a first-degree relative with mania, those who endorsed trauma and/or maltreatment, and individuals who reported high family conflict were significantly more likely to be assigned to the High/Variable and Moderate Symptom trajectories than the Low Symptoms trajectory. Our work is consistent with prior research showing a 14-fold increased risk of bipolar disorder in individuals with a first-degree relative affected by the condition (Mortensen et al., 2003). In our analysis, individuals in the High/Variable Symptoms trajectory had an almost three-fold risk of reporting a first-degree relative with bipolar disorder compared to those in the Low Symptom class. We also identified trauma and maltreatment as significant risk factors for mania symptoms, consistent with previous research linking childhood trauma to bipolar disorder (Etain et al., 2010, 2013; Watson et al., 2014). Childhood maltreatment has also been associated with more severe manic symptoms, earlier onset, rapid cycling, increased manic episodes, and a higher risk of transitioning from major depression to bipolar disorder (Agnew-Blais & Danese, 2016; Gilman et al., 2012). Our study also identified family conflict as a significant risk factor for High/Variable and Moderate Symptoms trajectories (relative to the Low Symptoms trajectory), consistent with previous findings linking family conflict to increased mania symptom severity (Sullivan et al., 2012). Our findings highlight the significant influence of family and environmental factors in the development and progression of mania symptoms.

Our findings extend our understanding of predictors of mania trajectories in young adolescents. Combined, mental health risk factors outperformed physical health, cognition, and family and environmental risk factors in predicting trajectory membership, emphasizing the high comorbidity between mania symptoms and transdiagnostic psychopathology symptoms in young people (Findling et al., 2013). These results are consistent with previous work showing that comorbid psychopathology symptoms – particularly depressive and anxiety symptoms – are among the strongest risk factors for a persistent, non-remitting course of mania symptoms in a general population sample (Beekman et al., 2023). Persistent anxiety and depressive symptoms – in combination with severe mania symptoms – also differentiate youth who later develop a bipolar spectrum disorder from youth who do not, highlighting their importance in bipolar disorder risk prediction (Van Meter et al., 2021). Sociodemographic factors were also strongly associated with trajectory membership, outperforming cognitive and physical health factors. This association is likely driven by trajectory group differences in socioeconomic status, consistent with prior work in the ABCD sample highlighting socioeconomic status as a key predictor in differentiating trajectories of depressive symptoms (Xiang et al., 2022). Ultimately, integrating mental, physical, cognitive, family and environmental risk factors will enhance risk prediction beyond individual domains.

In the future, we plan to explore a broader range of potential mania risk factors, including neurobiological markers (Abé et al., 2023; Cotovio & Oliveira-Maia, 2022), genetic predispositions (Merola et al., 2024), and psychosocial factors such as peer relationships and healthcare access disparities (Prichett et al., 2024), to develop a more comprehensive understanding of mania symptom risk. To advance early intervention approaches, efforts to identify more modifiable risk factors, such as lifestyle, social or metabolic risk factors, should be prioritized. Finally, predictive modeling approaches, such as risk calculators (e.g., Hafeman et al., 2017; Van Meter et al., 2021) may offer promising tools to estimate individual-level risk for elevated mania symptom trajectories and facilitate targeted preventive efforts.

Our study is strengthened by using a diverse, representative general population sample. By focusing on late childhood and early adolescence, we captured children at the earliest stages of symptom emergence. This approach enhances the generalizability of our findings and extends previous work that focused exclusively on clinical or high-risk samples (Birmaher et al., 2014; Findling et al., 2010; Mignogna & Goes, 2024; Weintraub, Schneck, Axelson, et al., 2020; Weintraub, Schneck, Walshaw, et al., 2020). By examining multiple risk factors across diverse domains, we were able to determine their relative importance in predicting mania symptom trajectories, as well as determine their combined utility in predicting trajectory group membership.

There are limitations associated with our study. Our study relied on parent-reported assessments of mania and mental health symptoms, which may be influenced by social desirability, parent-adolescent relationship dynamics, and cultural norms (Rescorla, 2016; Youngstrom et al., 2015). To enhance measurement validity, future research should incorporate youth self-report measures. At the time of analysis, data was only available for three data points per participant, limiting the ability to capture more complex, non-linear developmental patterns. However, youth in the ABCD cohort are continuing to receive annual assessments, which will provide insight into the longer-term course of mania symptoms through mid-to-late adolescence. While our final model entropy was less than ideal (0.71), our bias-adjusted approach mitigates the risk of class misclassification influencing associations between mania symptom trajectories and risk factors (Asparouhov & Muthén, 2014). We were not able to assess some established mania risk factors, such as goal attainment and negative life events, as these were not included in the baseline assessment. We modeled mania symptoms as a single construct; future work could seek to examine the distinct subconstructs of mania (e.g. pure, aggressive, psychotic, and mixed; Sato et al., 2002). The use of annual assessments, while valuable for capturing longitudinal trends, likely miss episodic symptom fluctuations; more frequent assessments and broader diagnostic frameworks (e.g. multi-informant assessments) could improve the precision of symptom tracking. Finally, because mania symptoms frequently co-occur with depressive symptoms, future research could examine both manic and depressive symptom trajectories concurrently to explore their interplay over time.

## Conclusion

This study identified three distinct trajectories of mania symptoms in a diverse community sample of over 10,000 U.S. adolescents. More severe symptom trajectories were associated with a broad array of risk factors, including increased mental health symptoms, physical comorbidities, and family and environmental factors. These findings highlight the importance of examining a range of risk factors – mental, physical, cognitive, family and environmental – to enhance risk prediction efforts in young people, and underscore the need for additional, developmentally-specific risk factors to improve early identification and intervention efforts in psychiatry.

## Data Availability

Data used in the preparation of this article were obtained from the Adolescent Brain Cognitive Development (ABCD) Study (https://abcdstudy.org), held in the NIMH Data Archive (NDA).

http://dx.doi.org/10.15154/z563-zd24

## Acknowledgements

This research was not preregistered. The authors declare no competing interests. RC was supported by the Tommy Fuss Center for Neuropsychiatric Disease Research Fellowship Award. MJ was supported by the National Institutes of Mental Health (R01MH129636) and the Tommy Fuss Center for Neuropsychiatric Research Next Generation Award.

Data used in the preparation of this article were obtained from the Adolescent Brain Cognitive Development (ABCD) Study (https://abcdstudy.org), held in the NIMH Data Archive (NDA). This is a multisite, longitudinal study designed to recruit more than 10,000 children aged 9-10 and follow them over 10 years into early adulthood. The ABCD Study® is supported by the National Institutes of Health and additional federal partners under award numbers U01DA041048, U01DA050989, U01DA051016, U01DA041022, U01DA051018, U01DA051037, U01DA050987, U01DA041174, U01DA041106, U01DA041117, U01DA041028, U01DA041134, U01DA050988, U01DA051039, U01DA041156, U01DA041025, U01DA041120, U01DA051038, U01DA041148, U01DA041093, U01DA041089, U24DA041123, U24DA041147. A full list of supporters is available at https://abcdstudy.org/federal-partners.html. A listing of participating sites and a complete listing of the study investigators can be found at https://abcdstudy.org/consortium_members/. ABCD consortium investigators designed and implemented the study and/or provided data but did not necessarily participate in the analysis or writing of this report. This manuscript reflects the views of the authors and may not reflect the opinions or views of the NIH or ABCD consortium investigators. The ABCD data repository grows and changes over time. The ABCD data used in this report came from ABCD Release 5.1 (DOI http://dx.doi.org/10.15154/z563-zd24).

