## Supplement for "Risk Factors Associated with Longitudinal Trajectories of Mania Symptoms in Youth"

### Supplemental Methods

*Mania symptoms*

We selected the parent-reported Parent General Behavior Inventory 10-item Mania Scale (PGBI-10M) to assess mania symptoms. While mania symptoms assessed via self-report are optimal, to our knowledge, no self-report mania scales were collected longitudinally and across successive time-points (see Supplemental Table 1). Among parent-reported questionnaires, we selected the PGBI due to its high validity, discrimination ability (Youngstrom et al., 2008), and popularity in previous ABCD studies (Farahdel et al., 2021; Palmer et al., 2020).

### Supplemental Table 1. Mania symptom questionnaires collected within the ABCD study

| Scale | Reporter | N items | *Assessed at* | | | | |
| --- | --- | --- | --- | --- | --- | --- | --- |
|  |  |  | Y0 | Y1 | Y2 | Y3 | Y4 |
| 7 Up Inventory (Youngstrom et al., 2013) | Youth | 7 | Y | Y | - | Y | - |
| Kiddie Schedule for Affective Disorders and Schizophrenia (K-SADS) –Screener questions for Bipolar Disorder (Kaufman et al., 1997) | Youth | 5 | Y | - | Y | - | - |
| KSADS – Bipolar Disorder module (Kaufman et al., 1997) | Youth | 90 | Y | - | Y | - | - |
| KSADS – Screener questions for Bipolar Disorder (Kaufman et al., 1997) | Parent | 5 | Y | - | Y | - | - |
| KSADS – Bipolar Disorder module (Kaufman et al., 1997) | Parent | 90 | Y | - | Y | - | - |
| Child Behavior Checklist (CBCL) – Mania symptoms subscale (Papachristou et al., 2013) | Parent | 19 | Y | Y | Y | Y | Y |
| Parent General Behavior Inventory (PGBI-10M) 10-item Mania Scale (Youngstrom et al., 2008) | Parent | 10 | Y | Y | Y | - | Y |

*Note.* Y indicates assessment completed; - indicates not assessed.

### Supplemental Table 2. Summary of studies from a literature review on risk factors for mania and manic symptoms in bipolar disorder

| Study | Risk factor examined | Design | N (participants/ studies), ages | Summary of main findings |
| --- | --- | --- | --- | --- |
| Physical Health | | | | |
| Jackson et al., 2003 | Sleep disturbances | Systematic review | 17 studies including 1191 participants | Among individuals with BD, sleep disturbance is the most frequently reported prodromal symptom of mania, with a median prevalence of 77%. |
| Melo et al., 2016 | Sleep disturbances | Systematic review | 30 studies including 7451 participants | BD is associated with disturbed sleep patterns, including inconsistent sleep/wake cycles, poor sleep quality, frequent nighttime awakenings, disrupted circadian rhythms, and elevated cortisol levels throughout the day. |
| Henquet et al., 2006 | Cannabis | Prospective cohort | 4815 participants (general population) aged 18- 64 years | Baseline cannabis use was associated with an increased risk of developing manic symptoms during follow-up after 3 years (adjusted OR = 2.70, 95% CI 1.54–4.75), after adjusting for age, sex, education level, ethnicity, marital status, neuroticism, use of other drugs, alcohol consumption, and the presence of depressive and manic symptoms at baseline. |
| van Laar et al., 2007 | Cannabis | Prospective cohort | 7076 participants at baseline; 4848 participants at follow-up (general population) aged 18-64 years | After adjusting for confounders, baseline cannabis use predicted a higher risk of experiencing a first bipolar episode (OR = 4.98; 95% CI 1.80–13.81) at 3-year follow-up. |
| Tijssen et al., 2010 | Cannabis | Prospective cohort | 1395 participants (general population) aged 14–17 years | In a general population cohort, cannabis use predicted the onset of manic symptoms (OR = 4.26, 95% CI 1.42–12.76; p < 0.01) at 8-year follow-up. |
| Baethge et al., 2008 | Cannabis | Prospective cohort | 166 participants with BD aged ≥18 years | In people with BD, cannabis use in both the preceding quarter (3 months) and in the same quarter is significantly associated with manic or hypomanic symptoms (preceding: regression coefficients [RC] = 0.111; 95% CI = 0.054–0.168; z = 3.80, p < 0.001; current: RC = 0.116; 95% CI = 0.053–0.178; z = 3.63, p < 0.001) using prospective follow-up study design (4.7 years). |
| Feingold et al., 2015 | Cannabis | Prospective cohort | 43,093 participants at baseline; 34,653 at follow-up (general population) aged ≥18 years | Weekly-to-daily use of cannabis was linked to a higher likelihood of developing BD in a 3-year prospective study (adjusted OR = 2.47, 95% CI = 1.03–5.92). However, daily use alone did not show a significant association (adjusted OR = 0.52, 95% CI 0.17–1.55). |
| Marwaha et al., 2018 | Cannabis | Prospective cohort | 3370 participants (general population) aged 17 years | Cannabis use at least two to three times per week was linked to an increased likelihood of developing hypomania at follow-up (up to 6 years; adjusted OR = 2.21, 95% CI = 1.49–3.28). A dose–response relationship was observed, with more frequent use showing stronger associations (any use versus weekly use). Cannabis use was found to mediate the relationship between childhood sexual abuse and hypomania, as well as between male sex and hypomania. |
| Denissoff et al., 2022 | Cannabis | Observational Study | 6325 participants (general population) aged age 15-16 years | Adolescent cannabis use was significantly associated with an increased risk of BD at follow-up (up to 18 years; HR = 3.46, 95% CI = 1.81–6.61). This association persisted after controlling for sex, family structure, parental psychiatric disorders and adolescent emotional and behavioral problems (HR = 2.34, 95% CI = 1.11–4.94). |
| Liu et al., 2015 | IBS | Retrospective cohort | 30,796 IBS-patients and 30,796 matched patients without IBS aged ≥20 years | The incidence of BD was higher in IBS patients compared to the matched cohort (incidence rate ratio = 2.63, 95% CI = 2.10–3.31, p < .001) using a 11-year follow-up design. |
| Tseng et al., 2016 | IBS | Meta-analysis | 6 studies including 177,117 IBS patients and 192,092 control subjects | The prevalence BD was significantly higher in patients with IBS compared to controls (OR = 2.48, 95% CI = 2.35–2.61, p < 0.001). |
| Liang & Chikritzhs, 2013 | Asthma | Cross-sectional | 8841 participants (general population) aged 18-85 years | Participants with a history of asthma lasting ≥6 months (but not asthma lasting <6 months) were at an increased risk of developing BD (Incidence Rate Ratio = 1.56, CI = 1.10-2.21), mania and hypomania (Incidence Rate Ratio = 1.71, CI = 1.20-2.43). |
| Liu et al., 2015 | Asthma | Retrospective cohort | 30,796 IBS-patients and 30,796 matched patients without IBS aged ≥20 years | Asthma is an independent risk factor for the development of BD in IBS patients (HR = 1.45, 95% CI = 1.08–1.95, p = .013) using a 11-year follow-up design. |
| Wu et al., 2016 | Asthma | Meta-analysis | 4 studies including 50,358 patients with asthma and 109,218 healthy controls | The prevalence of BD was significantly higher in asthmatic patients compared to healthy controls (OR = 2.12, 95% CI = 1.57–2.87, p < 0.001). |
| Wei et al., 2016 | Atopic disease | Prospective cohort | 49,804 participants (5075 with atopic diseases and 44,729 controls) aged 10-17 years | Adolescents with atopic diseases have an increased risk of developing BD (HR = 2.51, 95% CI = 1.71–3.67) compared to the general population at 10-year follow-up, with a dose-dependent relationship between having a greater number of atopic comorbidities and a greater likelihood of BD (1 atopic disease: HR = 1.40, 95% CI = 0.57–3.44; 2 atopic comorbidities: HR = 2.81, 95% CI = 1.68–4.68; ≥3 atopic comorbidities: HR = 3.02, 95% CI = 1.69–5.38). |
| Calkin et al., 2009 | Obesity | Cross-sectional | 276 participants with BD-I, BD-II, or BD-NOS aged 16-83 years | Higher BMI was associated with worse outcomes in BD, including a chronic course (p < 0.001), longer duration of illness (p = 0.02), lower scores on the Global Assessment of Functioning Scale (p = 0.02), greater disability (p = 0.002), more comorbid metabolic disorders (p=.001) and subthreshold anxiety disorders (p=.05). Patients in remission with lithium had lower BMIs (p = 0.01). |
| Kim et al., 2009 | Obesity | Cross-sectional | 184 patients with BD-I aged 19-64 years | 41.1% of obese patients showing depressive or mixed moods compared to 25.7% of non-obese patients (p=0.04). Obesity is more likely in those with depressive or mixed states than in those with manic states (p = 0.04). |
| Goldstein et al., 2011 | Obesity | Cross-sectional | 43,093 participants (general population) aged ≥18 years (adolescents 18–24 overrepresented) | The prevalence of obesity is significantly higher among individuals with BD compared to controls (AOR = 1.65, 95% CI = 1.45–1.89). |
| Zhao et al., 2016 | Obesity | Meta-analysis | 9 studies including 12,259 BD patients and 615,490 non-BD controls | Obesity is linked to a higher prevalence of BD (OR = 1.77, 95% CI = 1.40–2.23, p < 0.001). |
| Petri et al., 2017 | Obesity | Cross-sectional | 493 adult patients (2291 non-obese and 493 obese patients with MDD) | Obese patients with MDD experience more (hypo)manic switches during antidepressant treatment compared to non-obese MDD-patients (OR = 1.99, 95% CI = 1.57–2.54, p < .001). |
| Giménez-Palomo et al., 2022 | Obesity | Systematic review | 36 studies with 66,648 participants including both adolescents and adults | Obesity may be a risk factor for poorer outcomes in BD, including a chronic course, reduced global functioning, and rapid cycling. Obesity could potentially serve as a predictor of rapid cycling in BD. |
| Najar et al., 2024 | Obesity | Prospective cohort | 94,021 adult participants (22,127 BD patients and 71,894 controls) | Individuals with BD had higher BMI percentiles compared to controls. At the 50th percentile, BMI was 1.1 points higher in men 1 (95% CI=0.8-1.14, p < 0.001) points higher in women (95% CI=1.5-2.1, p < 0.001). At the 85th BMI percentile the gap was widest (men, 2.3 points, 95% CI =1.8-2.8, p < 0.001; women, 4.1 points 95% CI = 3.7-4.6, p < 0.001). BMI increased more rapidly over time in individuals with BD over a period of 12 years. The mean BMI increased by 1.1 points per decade for men with BD (95% CI: 0.7–1.4, p < 0.001), compared to 0.4 points in the general male population (95% CI: 0.3–0.5, p < 0.001). For women with BD, the mean BMI increased by 1.4 points per decade (95% CI: 1.1–1.7, p < 0.001), compared to 0.6 for women in the general population (95% CI: 0.5–0.7, p < 0.001). |
| Saunders et al., 2014 | Migraine | Retrospective cohort | 569 participants (412 BD, 157 healthy controls) aged ≥18 years | Migraine is significantly more prevalent in individuals with BD (31%) compared to healthy controls (6%). The risk of migraine is higher in women with BD compared to men (OR = 3.5; 95% CI, 2.1-5.8). Migraine is strongly linked to BD-II (OR = 9.9; 95% CI, 2.3-41.9) and mixed symptoms (OR = 3.5; 95% CI, 1.0-11.9). The presence of migraine is associated with an earlier onset of BD by two years, more severe depressive episodes (β = .13, p = .03), and increased frequency of depression at 5-year follow-up (β = .13, p = .03). |
| Fornaro & Stubbs, 2015 | Migraine | Meta-analysis | 14 studies including 3976 BD-patients (2161 BD-I, 647 BD-II), mean age 35.5 years (SD 7.6) | The overall pooled prevalence of migraine in BD was 34.8% (95% CI 25.54–44.69). The prevalence of migraine was higher among people with BD-II (54.17%, 95% CI = 31.52-75.95, n = 742) compared to BD-I (32.7%, 95% CI = 18.16-49.19, n = 2138, z = 3.97, p < .001). |
| Leo & Singh, 2016 | Migraine | Systematic review | 11 cross-sectional studies, aged ≥18 years, participants with BD and migraine | The weighted mean prevalence of BD in individuals with migraines was 5 to 9%, a 2.1-to-3.2-fold increase compared to the general population's 12-month prevalence rates. |
| Carta et al., 2014 | Multiple sclerosis | Cross-sectional | 1005 participants (201 MS patients and 804 controls) | Compared with controls, multiple sclerosis patients had a higher lifetime prevalence of BD-I (p = 0.05), BD-II (p < .001) and cyclothymia (p < .001). Among those with multiple sclerosis, bipolar spectrum disorders are more common than MDD when compared to the control group (p < 0.01). |
| Meier et al., 2020 | Multiple sclerosis | Record-linkage | ~15 million participants (128,194 MS-patients, 203,592 BD-patients, ~15 million control) | There is a significant increase in the risk of BD following hospital admission for multiple sclerosis (adjusted HR = 1.14, 95% CI = 1.04–1.24); conversely, the risk of developing MS after hospital admission for BD is significantly higher (adjusted HR = 1.73, 95% CI = 1.57–1.91) – however only when considering any diagnosis on the hospital record during 17 years of follow-up. |
| Liu et al., 2015 | Autoimmune diseases | Retrospective cohort | 30,796 IBS-patients and 30,796 matched patients without IBS aged ≥20 years | Autoimmune diseases are an independent risk factor for the development of BD in IBS patients (HR 1.52, 95% CI 1.07–2.17, p = .02) using a 11-year follow-up design. |
| Bachen et al., 2009 | Autoimmune diseases (systemic lupus) | Cross-sectional | 326 white women, mean age 47.9 years (SD = 11.3) | In individuals with systemic lupus erythematosus, the prevalence of BD-I is significantly higher than in the general population (SLE, 5.81% [95% CI: 3.3–8.4]; general population, 1.0% [95% CI 0.5-1.4], p < 0.001). |
| Cremaschi et al., 2017 | Autoimmune diseases | Cross-sectional | 10,283 participants (1952 BD-I, 1846 BD-II, 6485 controls), aged ≥18 years | Patients with BD experience higher rates of hypothyroidism not related to lithium (adjusted OR = 1.4, 95% CI = 1.1–1.7, p < 0.001), rheumatoid arthritis (adjusted OR = 17.9, 95% CI = 7.0–45.5, p < 0.001), and polymyalgia rheumatica (adjusted OR = 37.1, 95% CI = 4.8–289.3, p < 0.001) than healthy controls. Controls were more likely to experience systemic lupus erythematosus (adjusted OR = 6.0, CI = 2.9–12.5, p < 0.001). |
| Wang et al., 2018 | Autoimmune diseases | Retrospective cohort | 327,490 participants (65,498 patients with systemic autoimmune disease and 261,992 age-matched controls) | People with systemic autoimmune diseases have nearly double the incidence rate of BD compared to the control group (6.35 vs. 3.43 per 10,000 person-years; HR = 1.98, p < 0.0001). |
| Chen et al., 2021 | Autoimmune diseases | Systematic review and meta-analysis | 10 studies, adolescents and adults | The incidence of BD is significantly higher in patients with autoimmune disease compared to those without (mean difference = 1.54, 95% CI = 1.28–1.86, p < 0.00001). |
| Pugliese et al., 2019 | Pregnancy / birth (head circumference) | Observational study | 159 participants (74 patients with BD, 85 controls) aged 18–65 years | Head circumference < 32cm after birth, collected from medical records, was positively associated with a later BD diagnosis (χ2 = 22.331; p < 0.001). |
| Shintani et al., 2023 | Pregnancy / birth (multiple complications) | Systematic review and meta-analysis | 27 studies, individuals aged ≥18 years | Peripartum asphyxia (OR = 1.46, 95% CI = 1.02–2.11), maternal stress during pregnancy (OR = 12.00, 95% CI = 3.30–43.59), obstetric complications (OR = 1.41, 95% CI = 1.18–1.69), and birth weight below 2500 g (OR = 1.28, 95% CI = 1.04–1.56) are associated with an increased risk of developing BD. |
| Talati et al., 2013 | Pregnancy / birth (smoking) | Prospective cohort | 733 participants (79 BD, 654 matched controls) age ranging from the prenatal stage | After accounting for potential confounding factors, children exposed to maternal smoking during pregnancy showed a twofold increased risk of developing BD (OR = 2.014, 95% CI = 1.48–2.53, p = 0.01) within a maximum follow-up period of approximately 50 years. |
| Mackay et al., 2017 | Pregnancy / birth (smoking) | Prospective cohort | 2957 participants from birth, hypomania assessment aged 22-23 years | Smoking during pregnancy is not associated with lifetime hypomania in young adults. No significant associations were found with paternal smoking (p = 0.34) or early childhood exposure to environmental tobacco smoke (p = 0.26). |
| Chudal et al., 2015 | Pregnancy / birth (smoking) | Cross-sectional | 724 children diagnosed and/or treated with BD; 1419 matched controls | In unadjusted analyses, smoking during pregnancy was associated with a 1.41-fold increased risk of BD (OR = 1.41, 95% CI 1.12–1.79, p = 0.004), however when adjusting for potential covariates, the risk was not significant (OR = 1.14, 95% CI 0.88–1.49, p = 0.323). |
| Quinn et al., 2017 | Pregnancy / birth (smoking) | Prospective cohort | 1.7 million participants | Moderate smoking during pregnancy is associated with a 29% higher rate of BD in exposed offspring (HR = 1.29, 95% CI = 1.22-1.36), and high smoking with a 54% higher rate (HR = 1.54, 95% CI = 1.45-1.63). However, these associations are nonsignificant in sibling comparisons with within-family covariates in both moderate (HR = 1.06, 95% CI = 0.88-1.28) and high smoking (HR = 1.15, 95% CI = 0.92-1.44). |
| Mental Health | | | | |
| Brancati et al., 2021 | ADHD | Systematic review and meta-analysis | 10 studies including 1248 participants diagnosed with ADHD aged 6-25 years | The likelihood of BD occurrence is approximately 10 times higher in individuals with ADHD compared to healthy controls (OR = 10.30, 95% CI = 4.74 - 22.38, p < 0.0001). Prospectively, individuals with ADHD are approximately 9 times more likely to develop BD compared to healthy controls (Relative Risk [RR] = 8.97, 95% CI = 4.26 - 18.87 p < 0.0001). |
| Biederman et al., 2009 | ADHD | Retrospective cohort | 832 participants (280 with ADHD, 74 ADHD-siblings, 242 controls, 236 control-siblings) aged 6–18 years | ADHD increases the risk of transitioning from unipolar depression to BD (28% compared to 6% in healthy control; z = 2.80, p = 0.005). |
| Brus et al., 2014 | ADHD | Narrative review | NR | Around 20% of adults with ADHD also experience BD, while 10% to 20% of individuals with BD have adult ADHD. The coexistence of BD and ADHD is linked to an earlier onset, a more persistent and disabling progression of BD, and a higher likelihood of additional psychiatric conditions. |
| Sandstrom et al., 2021 | ADHD | Systematic review and meta-analysis | 92 studies including 17,089 people with BD | ADHD is three times more common in people with mood disorders compared to those without (RR = 3.42, 95% CI = 2.81–4.16, p < 0.001, I^2^ = 59.70%) and 1.7 times more common in BD compared to MDD (RR = 1.72, 95% CI = 1.20–2.47, p =  0.003). |
| Morcillo et al., 2012 | Behavioral problems / conduct disorder | Cross-sectional | 14,524 male participants (929 with conduct disorder, 13,595 controls) aged ≥18 years | Conduct disorder is associated with increased risk for BD (adjusted OR = 2.04, 95% CI = 1.62-2.57).  23.57% (SE = 1.54) of men with conduct disorder have BD, compared to 4.96% (SE = 0.21) in men without conduct disorder. |
| Tseng et al., 2015 | Behavioral problems / conduct disorder | Cross-sectional | 44 participants (23 at risk, due to a first-degree relative with BD, 21 controls) aged 3.45– 5.96 years | Children in the high-risk group experienced more problems in anger modulation (22% versus 0% in controls, p = .050) and more problems in in behavior regulation (39% versus 10% in controls; p = .036). |
| Verdolini et al., 2018 | Behavioral problems / conduct disorder | Systematic review and meta-analysis | 12 studies including 58,475 BD participants aged ≥18 years | The prevalence of violent criminal behavior in individuals with BD was found to be 7.1% (95% CI, 3.0‒16.5), The association between BD and violent criminal behavior compared to the general population was significant (OR = 5.22; 95% CI, 1.34‒20.25; p < .001), however, in sensitivity analyses, this association was no longer significant (OR = 2.78; 95% CI, 0.69‒11.29, p = .15). |
| Vaughn et al., 2010 | Behavioral problems / conduct disorder | Cross-sectional | 43,093 men (general population) aged 18-34 years | Individuals with a history of bullying others had a significantly higher likelihood of lifetime BD (OR = 1.47, 95% CI = 1.20–1.80). |
| Kennedy et al., 2005 | Behavioral problems / conduct disorder | Retrospective cohort | 246 participants with BD-I aged ≥16 years | In individuals with BD-I, younger age of onset was associated with childhood antisocial behavior (F = 4.3, p = 0.04) and male gender (F = 3.9, p < 0.05) |
| Carlson et al., 2000 | Behavioral problems / conduct disorder | Prospective cohort | 53 hospitalized participants with BD (23 early-onset, illness emerged <21 years and 30 later-onset illness emerged >30 years) | Subjects with early-onset psychotic mania were significantly more likely to have had clinically significant childhood behavior disorders, reporting a higher mean number of conduct disorder symptoms than adult-onset cases (mean = 2.13 (SD = 1.13) vs. 0.78 (SD = 1.12); p < 0.0001). Over the 24-month follow-up, manic episode recurrence was also substantially more frequent in the early-onset group (64.7% vs. 12.5%; p < 0.01). |
| Buckley et al., 2023 | Anxiety disorders | Systematic review | 16 studies including 2,433,761 participants from 10 countries; primarily children and adolescents with anxiety disorders | Anxiety disorders in childhood and adolescence significantly increase the risk of developing BD later in life with ORs ranging from approximately 4.5 to 12 depending on the type of anxiety disorder and study population. |
| Johnson et al., 2000 | Anxiety disorders | Prospective cohort | 717 youth participants, mean age of 14 years at baseline | Adolescents with anxiety disorders are significantly more likely to develop BD or clinically significant manic symptoms by early adulthood within a 10-year follow-up compared to other adolescents without anxiety disorders in a representative community sample (OR = 4.69, 95% CI = 1.78–12.38, p < 0.01). Adolescents with manic symptoms are at increased risk for developing anxiety disorders (adjusted OR = 1.38, 95% CI = 1.15–1.66) and depressive disorders (adjusted OR = 1.27, 95% CI = 1.02–1.59) in early adulthood, even after controlling for adolescent diagnoses. |
| Gilman et al., 2012 | Anxiety disorders | Prospective cohort | 6214 cases of MDD aged ≥18 years | In individuals with MDD, history of social phobia (OR = 2.20, 95% CI = 1.47–3.30) and generalized anxiety disorder (OR = 1.58, 95% CI = 1.06–2.35) is associated with the transition to BD within 3 years of follow-up. |
| Katz et al., 2021 | Behavioral activation system | Meta-analysis | 23 studies with self-reported (hypo)mania (general population); 33 studies with diagnosed BD-patients and controls | Behavioral activation system (BAS) sensitivity was associated with higher (hypo)manic risk in the general population (g = 0.74, 95% CI 0.54-0.93), while no significant relationship was found with behavioral inhibition system (BIS) sensitivity (g = -0.08, 95% CI = − 0.28-0.12). Euthymic individuals with BD had higher BAS sensitivity (g = 0.20, 95% CI = 0.06-0.33) and BIS sensitivity (g = 0.64, 95% CI = 0.47-0.81) than healthy controls. |
| Axelson et al., 2011 | Psychiatric hospitalizations | Prospective cohort | 140 participants with BD-NOS aged 7-17 years | Previous psychiatric hospitalization is significantly associated with the conversion from BD-NOS to BD-I or BD-II in youth (HR = 2.48, 95% CI 1.43–4.32, p = 0.001) over five years. |
| Cognitive Functioning | | | | |
| Kurtz & Gerraty, 2009 | Neurocognition / verbal learning / verbal memory | Meta-analysis | 60 studies (42 studies including 1,197 BD patients in euthymia; 13 studies including 314 patients in manic/ mixed state; 5 studies including 96 patients in depressed state) | In individuals with BD, euthymic phases are associated with large effect-size impairments in verbal learning (d = .81) and delayed verbal and non-verbal memory (d = .80–.92) compared to controls. In manic/mixed state, impairments are exaggerated, especially in verbal learning (d = 1.43), delayed free recall (d = 1.05), and executive function (d = .64–.72) and attention (d = .79). |
| Miskowiak et al., 2022 | General cognitive impairment | Systematic review | 12 studies in individuals with BD (36-76 participants); 7 studies in at-risk individuals (84-234 participants) | In individuals with BD, general cognitive impairment, poorer verbal memory, executive function, and positive bias were associated with subsequent hypomanic or manic relapses. In at-risk first-degree relatives, impairments in attention, verbal memory, and executive functions, along with a positive bias, were linked to subsequent illness onset. |
| Sankar et al., 2023 | Verbal memory | Prospective cohort | 518 participants (438 BD, 80 MDD) | Verbal memory impairments (z-score ≤ -1) are significantly associated with an increased risk of future psychiatric hospitalization in individuals with BD (HR = 1.84, 95% CI: 1.05–3.25, p = 0.034), even after adjusting for illness duration, at up to 11 years of follow-up. |
| Lex et al., 2017 | Verbal memory | Cross-sectional | 165 participants (57 controls, 45 at risk for depression, and 63 at risk for mania); adolescents and young adults | No overall verbal memory deficits were observed across groups (p = 0.92). |
| Fleck et al., 2003 | Verbal memory (recall, recognition) | Cross-sectional | 68 participants (8 manic hospitalized patients, 6 mixed hospitalized patients, 14 euthymic outpatients, 40 healthy controls) | Both manic and euthymic patient groups performed significantly worse than healthy controls on all recall measures (p < 0.05 for all comparisons). Manic patients also performed worse than both healthy subjects and euthymic patients in recognition (p < 0.05). |
| Family and Environmental Factors | | | | |
| Etain et al., 2010 | Childhood trauma (multiple trauma, emotional abuse) | Cross-sectional | 206 BD patients (155 BD-I and 51BD-II) and 94 control subjects | Individuals with BD reported greater childhood trauma than controls (mean 43.4 [SD 13.5] vs 36.3 [SD 7.4], p<.001). Multiple trauma (at least two subtypes at low intensity) was significantly more frequent in patients with BD (63%) than controls (33%; OR = 3.48, 95% CI = 2.08–5.82). The cumulative number of traumatic events was strongly associated with BD status (z = −4.52, p < .001). Emotional abuse was a significant predictor of BD (OR = 2.14, 95% CI = 1.51–3.02, p < .001). |
| Etain et al., 2013 | Childhood trauma (maltreatment / abuse) | Cross-sectional | 587 adults with BD | Both emotional and sexual abuse independently predicted an earlier age of onset (p = .002 for each). Sexual abuse was the strongest predictor of rapid cycling symptoms (OR = 2.04, 95% CI = 1.21- 3.42, P = .007). |
| Watson et al., 2014 | Childhood trauma (maltreatment/ abuse), emotional neglect | Cross-sectional | 115 participants (31 outpatients with BD-I, 25 outpatients with BD-II, 4 outpatients with BD-NOS, 55 controls), aged 18–65 years | The overall amount of childhood trauma is significantly associated with BD (β = 0.08, p = 0.001), with emotional neglect being most relevant trauma type for distinguishing between BD and control participants (β = 0.185, p < 0.001). More specifically, the Childhood Trauma Questionnaire (CTQ) total score in the BD group was significantly higher than in healthy controls (Mann–Whitney U = 490.0, p < 0.001), with significant differences across emotional abuse, physical abuse, and physical neglect (all p<.001). Sexual abuse showed no significant difference between the groups (p = 0.131). |
| Agnew-Blais & Danese, 2016 | Childhood trauma (maltreatment / abuse) | Meta-analysis | 30 studies with maltreatment occurring before the age of 18 | Patients with BD and a history of childhood maltreatment showed greater mania severity (OR = 2.02, 95% CI = 1.21–3.39, p = 0.008), earlier onset of BD (OR = 1.85, 95% CI = 1.43–2.40, p < 0.0001), higher risk of rapid cycling (OR = 1.89, 95% CI = 1.45–2.48, p < 0.0001), and a greater number of manic episodes (OR = 1.26, 95% CI = 1.09–1.47, p = 0.003) compared to those without childhood maltreatment. |
| Palmier-Claus et al., 2016 | Childhood trauma (maltreatment / abuse) | Meta-analysis | 19 studies with maltreatment occurring before the age of 19 | Individuals with BD were more likely to have experienced childhood adversity compared to non-clinical controls (OR = 2.63, 95% CI = 2.00–3.47, p < 0.001). Emotional abuse had the strongest association (OR = 4.04, 95% CI = 3.12–5.22), |
| Gilman et al., 2015 | Childhood trauma (maltreatment / abuse) | Prospective cohort | 33,375 participants (general population), including 811 with manic episode, aged ≥18 years | Factors that increased the risk of a first-onset manic episode are a history of childhood abuse (OR = 2.23; 95% CI = 1.71–2.91) and sexual maltreatment (OR = 2.10; 95% CI = 1.55–2.83) within 3 years of follow-up. |
| Gilman et al., 2012 | Childhood trauma (maltreatment / abuse) | Prospective cohort | 6214 cases of MDD aged ≥18 years | In individuals with MDD, history of child abuse predicted the transition to BD (OR = 1.26, 95% CI 1.12–1.42) within 3 years of follow-up. |
| Sullivan et al., 2012 | Childhood trauma (conflict) | Prospective cohort | 58 adolescents diagnosed with BD (38 with BD-I, 6 with BD-II, 14 with BD-NOS) and their families aged 12-17 years | Parent-reported family conflict at baseline predicted higher levels of mania symptoms over the 2-year study (low-conflict: mean 16.62, SD 12.00; high-conflict: mean 19.53, SD 10.67p = .04). Adolescents from high-conflict families had slower reductions in mania symptoms compared to those from low-conflict families (low-conflict: β = −0.77; high-conflict: −0.46; p < 0.0001). Decreases in family conflict as reported by parents were associated with decreases in manic symptoms (Fp=.02). |
| Gilman et al., 2012 | Problem with social support group | Prospective cohort | 6214 cases of MDD aged ≥18 years | Past-year problems with social support group predicted the transition to BD in patients with MDD (OR = 1.79, 95% CI 1.19–2.68) within 3 years of follow-up. |
| Mortensen et al., 2003 | Childhood trauma (maternal loss) | Prospective cohort | 2.1 million (general population), 2,299 BD patients including children, adolescents and adults | Parental loss before the fifth birthday is associated with an increased risk for BD (OR 4.05, 95% CI 1.68–9.77) |
| Mortensen et al., 2003 | First degree relative with bipolar disorder | Prospective cohort | 2.1 million (general population), 2,299 BD patients including children, adolescents and adults | A first-degree relative with BD is associated with increased risk for BD (OR 13.63, 95% CI = 11.81–15.71). |
| Axelson et al., 2011 | First and second degree relative with mania or hypomania | Prospective cohort | 140 participants with BD-NOS aged 7-17 years | A family history of mania or hypomania increases the likelihood of conversion from BD-NOS to BD-I or BD-II in youth (HR = 3.08, 95% CI = 1.77–5.38, p < 0.0001) over five years. |
| Johnson et al., 2000 | Goal-directed life events | Prospective cohort | 43 participants with BD-I aged 18-65 years | Goal-attainment life events are significantly associated with an increase in manic symptoms in the two months following the event (r = -0.37, p = .01). Goal-attainment events predict more manic symptom increases than general positive events (z = -1.95, p < 0.05). |
| Nusslock et al., 2007 | Goal-directed life events | Prospective cohort | 159 participants (68 with cyclothymia/ BD-II, 91 controls) aged 18-24 years | In individuals with BD, final exams (a goal-directed life event) were associated with an increase in hypomania (OR = 16.98, p = .05). |
| Lex et al., 2017 | Goal-directed and stressful life events | Meta-analysis | 42 studies including 36,524 participants (4562 with BD, 31,962 controls) | Patients with BD reported significantly more significant life events, including stress, daily hassles, or goal attainment, before an acute episode compared to their own healthy intervals (Hedges’ g = 0.51, SE = 0.13, 95%-CI = 0.26–0.76, p < .01). Patients with BD also reported significantly more life events prior to an acute mood episode than healthy individuals (Hedges' g = 0.57, SE = 0.12, 95%-CI = 0.36–0.81, p < .01). |
| Koenders et al., 2014 | Negative life events | Prospective cohort | 173 BD outpatients (121 BD-I, 52 BD-II, 2 BD-NOS, 1 cyclothymia); mean age 49.9 years (SD 11.4) | Negative events predicted an increase in mania symptoms at follow-up in BD-I patients (β = 230, SE = .063, p = .001). Positive events predicted an increase in mania symptoms at follow-up in BD-I patients (β = .329, SE = .081, p = .019). |
| Gilman et al., 2015 | Negative life events (personal loss) | Prospective cohort | 33,375 participants (general population, including 811 with manic episode) aged ≥18 years | Personal loss associated with increased risk for a first-onset manic episode (OR = 1.41; 95% CI = 1.02–1.94) within 3 years of follow-up. |
| Gilman et al., 2015 | Financial instability / economic hardship | Prospective cohort | 33,375 participants (general population; 811 with manic episode) aged ≥18 years | Financial and interpersonal instability (OR = 2.62; 95% CI = 1.95–3.52) and economic hardship (OR = 1.93; 95% CI = 1.36–2.73) associated with increased risk for first-onset manic episode within 3 years of follow-up. |
| Demographics | | | | |
| Gilman et al., 2012 | Race | Prospective cohort | 6,214 cases of MDD aged ≥18 years | In individuals with MDD, black individuals were more likely than white individuals to transition to BD (OR = 1.72, 95% CI = 1.04-2.84) within 3 years of follow-up. |
| Axelson et al., 2011 | Ethnicity | Prospective cohort | 140 participants with BD-NOS aged 7-17 years | Among individuals with BD-NOS, white youth were more likely to convert to BD-I or BD-II than non-white youth (HR = 2.70; 95% CI = 1.15–6.33, p = 0.02) over five-year follow-up. |
| Kennedy et al., 2005 | Gender | Retrospective cohort | 246 participants with BD-I aged ≥16 years (141 female, 105 male) | Male gender associated with earlier onset compared to female gender (30 years [SD 13.4] vs 35.1 years [SD 16.2], p=.007). |
| Dell’Osso et al., 2021 | Gender | Systematic review | 10 large sample studies including 47,878 BD patients | All ten large-sample studies (n > 1000) reported a predominance of female participants among BD patients. Female representation ranged from 57.4-65.0%. |
| Gilman et al., 2015 | Household income | Prospective cohort | 33,375 participants (811 with manic episode) aged ≥18 years | Economic disadvantage during childhood (OR = 1.27; 95% CI = 0.99–1.62) increases the risk of a first-onset manic episode within 3 years of follow-up. |

*Note.* BD, bipolar disorder; BD-I, bipolar I disorder; BD-II, bipolar II disorder; BD-NOS, bipolar disorder not otherwise specified; MDD, major depressive disorder; ADHD, attention deficit hyperactivity disorder; IBS, irritable bowel syndrome; BMI, body mass index; CI, confidence interval; HR, hazard ratio; OR, odds ratio; NR, not reported; SD, standard deviation.

### Supplemental Table 3. Overview of assessed items by risk factor category

| Assessed risk factor | High/Variable exclusions | Questionnaire | ABCD item code | Item text/instructions |
| --- | --- | --- | --- | --- |
| Physical Health | | | | |
| Sleep disturbances |  | Sleep disturbance Scale for Children (Total Score; Bruni et al., 1996) | sds_p_ss_total |  |
| Cannabis use | Low number of endorsements at baseline |  |  |  |
| Irritable bowel syndrome |  | Child Behavior Checklist (Achenbach, 1991) |  | Below is a list of items that describe children and youths. For each item, rate whether it describes your child now or within the past 6 months using the following scale: 0 = Not True, 1 = Somewhat/Sometimes True, 2 = Very True/Often True |
|  |  |  | cbcl_q56c_p | Nausea, feels sick |
|  |  |  | cbcl_q56f_p | Stomachaches |
|  |  |  | cbcl_q56g_p | Vomiting, throwing up |
| Asthma |  |  | medhx_2a | Since we last saw you, has she/he been to a doctor for any of these things? …Asthma |
| Atopic disease | No conceptually matching variable available |  |  |  |
| Obesity |  |  | anthroweight1lb, anthroweight2lb, anthroweight3lb, anthro_1_height_in, anthro_2_height_in, anthro_3_height_in | Weight in lb. Height in in. |
| Migraine | Low number of endorsements at baseline |  |  |  |
| Multiple sclerosis | No conceptually matching variable available |  |  |  |
| Autoimmune diseases | No conceptually matching variable available |  |  |  |
| Pregnancy complications |  | ABCD Developmental History Questionnaire (Barch et al., 2018) |  | During the pregnancy with this child, did you/biological mother have any of the following conditions? |
|  |  |  | devhx_10a3_p | Severe nausea and vomiting extending past the 6th month or accompanied by weight loss |
|  |  |  | devhx_10b3_p | Heavy bleeding requiring bed rest or special treatment |
|  |  |  | devhx_10c3_p | Pre-eclampsia, eclampsia, or toxemia |
|  |  |  | devhx_10d3_p | Severe gall bladder attack |
|  |  |  | devhx_10e3_p | Persistent proteinuria |
|  |  |  | devhx_10f3_p | Rubella (German measles) during first 3 months of pregnancy |
|  |  |  | devhx_10g3_p | Severe anemia |
|  |  |  | devhx_10h3_p | Urinary tract infections |
|  |  |  | devhx_10i3_p | Pregnancy-related diabetes |
|  |  |  | devhx_10j3_p | Pregnancy-related high blood pressure |
|  |  |  | devhx_10k3_p | Previa, abruptio, or other problems with the placenta |
|  |  |  | devhx_10l3_p | An accident or injury requiring medical care |
|  |  |  | devhx_10m3_p | Any other conditions requiring medical care |
| Smoking during pregnancy | Insufficient evidence (<2 studies) |  |  |  |
| Obstetric complications |  | ABCD Developmental History Questionnaire (Barch et al., 2018) |  | Did he/she have any of the following complications at birth? |
|  |  |  | devhx_14a3_p | Blue at birth |
|  |  |  | devhx_14b3_p | Slow heart beat |
|  |  |  | devhx_14c3_p | Did not breathe at first |
|  |  |  | devhx_14d3_p | Convulsions |
|  |  |  | devhx_14e3_p | Jaundice needing treatment |
|  |  |  | devhx_14f3_p | Required oxygen |
|  |  |  | devhx_14g3_p | Required blood transfusion |
|  |  |  | devhx_14h3_p | Rh incompatibility |
| Premature birth |  | ABCD Developmental History Questionnaire (Barch et al., 2018) | devhx_12a_p | Was the child born prematurely? |
| Cesarean section |  | ABCD Developmental History Questionnaire (Barch et al., 2018) | devhx_13_3_p | Was your child born by Caesarian section? |
| Mental Health | | | | |
| Depression |  | Child Behavior Checklist (Achenbach, 1991) | cbcl_scr_dsm5_depress_r |  |
| Attention deficit hyperactivity disorder |  | Child Behavior Checklist (Achenbach, 1991) | cbcl_scr_dsm5_adhd_r |  |
| Behavioral problems / conduct disorder |  | Child Behavior Checklist (Achenbach, 1991) | cbcl_scr_dsm5_conduct_r |  |
| Oppositional-defiant disorder |  | Child Behavior Checklist (Achenbach, 1991) | cbcl_scr_dsm5_opposit_r |  |
| Anxiety |  | Child Behavior Checklist (Achenbach, 1991) | cbcl_scr_dsm5_anxdisord_r |  |
| Psychiatric hospitalizations | Insufficient evidence (<2 studies) |  |  |  |
| Behavioral activation system | Insufficient evidence (<2 studies) |  |  |  |
| Cognitive Functioning | | | | |
| General cognition | Insufficient evidence (<2 studies) |  |  |  |
| Verbal learning impairment |  | Rey Auditory Verbal Learning Test (Schmidt, 1996) | Sum of pea_ravlt_sd_trial_i_tc: pea_ravlt_sd_trial_v_tc |  |
| Family and Environmental Factors | | | | |
| Childhood trauma |  | Kiddie Schedule for Affective Disorders and Schizophrenia - Post-Traumatic Stress Disorder (Kaufman et al., 1997) | ksads_ptsd_raw_754_p | A car accident in which your child or another person in the car was hurt bad enough to require medical attention |
|  |  |  | ksads_ptsd_raw_755_p | Another significant accident for which your child needed specialized and intensive medical treatment |
|  |  |  | ksads_ptsd_raw_756_p | Witnessed or caught in a fire that caused significant property damage or personal injury |
|  |  |  | ksads_ptsd_raw_757_p | Witnessed or caught in a natural disaster that caused significant property damage or personal injury |
|  |  |  | ksads_ptsd_raw_758_p | Witnessed or present during an act of terrorism (e.g., Boston marathon bombing) |
|  |  |  | ksads_ptsd_raw_759_p | Witnessed death or mass destruction in a war zone |
|  |  |  | ksads_ptsd_raw_760_p | Witnessed someone shot or stabbed in the community |
|  |  |  | ksads_ptsd_raw_761_p | Shot, stabbed, or beaten brutally by a non-family member |
|  |  |  | ksads_ptsd_raw_762_p | Shot, stabbed, or beaten brutally by a grown up in the home |
|  |  |  | ksads_ptsd_raw_763_p | Beaten to the point of having bruises by a grown up in the home |
|  |  |  | ksads_ptsd_raw_764_p | A non-family member threatened to kill your child |
|  |  |  | ksads_ptsd_raw_765_p | A family member threatened to kill your child |
|  |  |  | ksads_ptsd_raw_766_p | Witness the grownups in the home push, shove or hit one another |
|  |  |  | ksads_ptsd_raw_767_p | A grown up in the home touched your child in their privates, had your child touch their privates, or did other sexual things to your child |
|  |  |  | ksads_ptsd_raw_768_p | An adult outside your family touched your child in their privates, had your child touch their privates or did other sexual things to your child |
|  |  |  | ksads_ptsd_raw_769_p | A peer forced your child to do something sexually |
|  |  |  | ksads_ptsd_raw_770_p | Learned about the sudden unexpected death of a loved one |
| Childhood maltreatment |  | Kiddie Schedule for Affective Disorders and Schizophrenia - Post-Traumatic Stress Disorder (Kaufman et al., 1997) | ksads_ptsd_raw_761_p | Shot, stabbed, or beaten brutally by a non-family member |
|  |  |  | ksads_ptsd_raw_762_p | Shot, stabbed, or beaten brutally by a grown up in the home |
|  |  |  | ksads_ptsd_raw_763_p | Beaten to the point of having bruises by a grown up in the home |
|  |  |  | ksads_ptsd_raw_764_p | A non-family member threatened to kill your child |
|  |  |  | ksads_ptsd_raw_765_p | A family member threatened to kill your child |
|  |  |  | ksads_ptsd_raw_767_p | A grown up in the home touched your child in their privates, had your child touch their privates, or did other sexual things to your child |
|  |  |  | ksads_ptsd_raw_768_p | An adult outside your family touched your child in their privates, had your child touch their privates or did other sexual things to your child |
|  |  |  | ksads_ptsd_raw_769_p | A peer forced your child to do something sexually |
| Childhood maltreatment |  | Children’s Report of Parental Behavior Inventory (Schaefer, 1965) | Instructions / Rating | 1 = Not like him/her  2 = Somewhat like him/her  3 = A lot like him/her  (Recoded, “Not like him/her” = Neglect) |
|  |  |  | crpbi_parent1_y | First caregiver (caregiver participating in study/completing protocol). Makes me feel better after talking over my worries with him/her. |
|  |  |  | crpbi_parent2_y | First caregiver (caregiver participating in study/completing protocol). Smiles at me very often. |
|  |  |  | crpbi_parent3_y | First caregiver (caregiver participating in study/completing protocol). Is able to make me feel better when I am upset. |
|  |  |  | crpbi_parent4_y | First caregiver (caregiver participating in study/completing protocol). Believes in showing his/her love for me. |
|  |  |  | crpbi_parent5_y | First caregiver (caregiver participating in study/completing protocol). Is easy to talk to. |
| Problem with social support group | Insufficient evidence (<2 studies) |  |  |  |
| First-degree relative with mania |  | ABCD Family History Questionnaire (Barch et al., 2018) | famhx_ss_fath_prob_ma_p (father)  famhx_ss_moth_prob_ma_p (mother)  famhx_ss_fulsiby1_prob_ma_p (full sibling younger)  famhx_ss_fulsiby2_prob_ma_p  famhx_ss_fulsiby3_prob_ma_p  famhx_ss_fulsiby4_prob_ma_p  famhx_ss_fulsiby5_prob_ma_p  famhx_ss_fulsibo1_prob_ma_p (full sibling older)  famhx_ss_fulsibo2_prob_ma_p  famhx_ss_fulsibo3_prob_ma_p  famhx_ss_fulsibo4_prob_ma_p  famhx_ss_fulsibo5_prob_ma_p | Has ANY blood relative of your child ever had a period of time when others were concerned because they suddenly became more active day and night and seemed not to need any sleep and talked much more than usual for them? |
| Socioeconomic status |  | ABCD Parent Demographics Survey (Barch et al., 2018) | demo_income_v2 | How much did you earn, before taxes and other deductions, during the past 12 months? |
|  |  |  | demo_roster_v2 | How many people are living at your address? INCLUDE everyone who is living or staying at your address for more than 2 months. |
| Goal-directed life events | No conceptually matching variable available |  |  |  |
| Negative / stressful life events | No conceptually matching variable available |  |  |  |

### Supplemental Table 4. Results from Wald X^2^ tests testing equality of beta coefficients in predicting mania trajectory membership

| Coefficient 1 | Coefficient 2 | Wald Χ^2^ test value | *df* | *p*_FDR_ |
| --- | --- | --- | --- | --- |
| Mental Health | Family & Environment | 55.73 | 1 | **1.44e-13** |
| Mental Health | Physical Health | 48.61 | 1 | **5.30e-12** |
| Mental Health | Cognition | 274.99 | 1 | **2.00e-16** |
| Mental Health | Sociodemographics | 0.34 | 1 | 6.08e-01 |
| Sociodemographics | Cognition | 7.64 | 1 | **8.73e-03** |
| Sociodemographics | Family & Environment | 2.63 | 1 | 1.41e-01 |
| Sociodemographics | Physical Health | 5.72 | 1 | **2.43e-02** |
| Family & Environment | Cognition | 8.56 | 1 | **5.47e-03** |
| Family & Environment | Physical Health | 1.28 | 1 | 3.20e-01 |
| Cognition | Physical Health | 0.70 | 1 | 4.64e-01 |

All risk factors were included in a multivariable regression model predicting trajectory membership (latent variable), and pair of coefficients (coefficients 1 and 2) were compared using Wald tests of the linear hypothesis *H*_0_: β_1_ - β_2_ = 0, which follows a Ꭓ^2^ distribution with one degree of freedom. Values in bold are significant after correction for multiple comparisons (*p*_FDR_<.05). The Mental Health beta coefficient is the sum of coefficients for depression, oppositional defiant disorder, attention deficit hyperactivity disorder, conduct disorder, and anxiety. The Sociodemographic coefficient comprises the summed coefficients for sex, race, ethnicity, and socioeconomic status. The Family & Environment coefficient is the sum of coefficients for first-degree relatives with mania, trauma, maltreatment and family conflict. The Physical Health beta coefficient is the summed coefficients for irritable bowel syndrome, pregnancy complications, obstetric complications, asthma, sleep disturbances, body mass index, cesarean section, premature birth, and low birth weight. The Cognition beta coefficient is the coefficient for verbal learning impairment.

### Supplemental Table 5. Associations between mania trajectories and sleep disorder subscales

|  | High/Variable Symptoms  vs Low Symptoms | | | Moderate Symptoms  vs Low Symptoms | | | High/Variable Symptoms  vs Moderate Symptoms | | |
| --- | --- | --- | --- | --- | --- | --- | --- | --- | --- |
|  | aOR | 95% CI | *d* | aOR | 95% CI | *d* | aOR | 95% CI | *d* |
| Disorders of Initiating and Maintaining Sleep | 3.40*** | 2.97-3.90 | 0.67 | 2.06*** | 1.87-2.28 | 0.40 | 1.65*** | 1.46-1.86 | 0.28 |
| Sleep Breathing Disorders | 1.54*** | 1.45-1.63 | 0.24 | 1.22*** | 1.13-1.31 | 0.11 | 1.26*** | 1.20--1.34 | 0.13 |
| Disorder of Arousal | 2.02*** | 1.86-2.20 | 0.39 | 1.54*** | 1.40-1.71 | 0.24 | 1.31*** | 1.19-1.43 | 0.15 |
| Sleep-Wake Transition Disorders | 2.79*** | 2.53-3.09 | 0.57 | 1.76*** | 1.63-1.90 | 0.31 | 1.59*** | 1.43-1.76 | 0.25 |
| Sleep Hyperhydrosis | 1.69*** | 1.51-1.89 | 0.29 | 1.44*** | 1.29-1.60 | 0.20 | 1.18*** | 1.09-1.27 | 0.09 |
| Disorders of Excessive Somnolence | 3.21*** | 2.78-3.70 | 0.64 | 2.05*** | 1.86-2.26 | 0.40 | 1.56*** | 1.39-1.76 | 0.25 |

The reference category is the Low Symptoms trajectory. Adjusted odds ratios (aOR) are presented. All models are adjusted for sex, race, ethnicity, and socioeconomic status. *d*, Cohen’s d effect size. Statistical significance is denoted as: * = *p*_FDR_<0.05; *

* = *p*_FDR_<0.01; *** *p*_FDR_<0.001.

### Supplemental Figure 1. Distribution of mania symptom scores at each time point

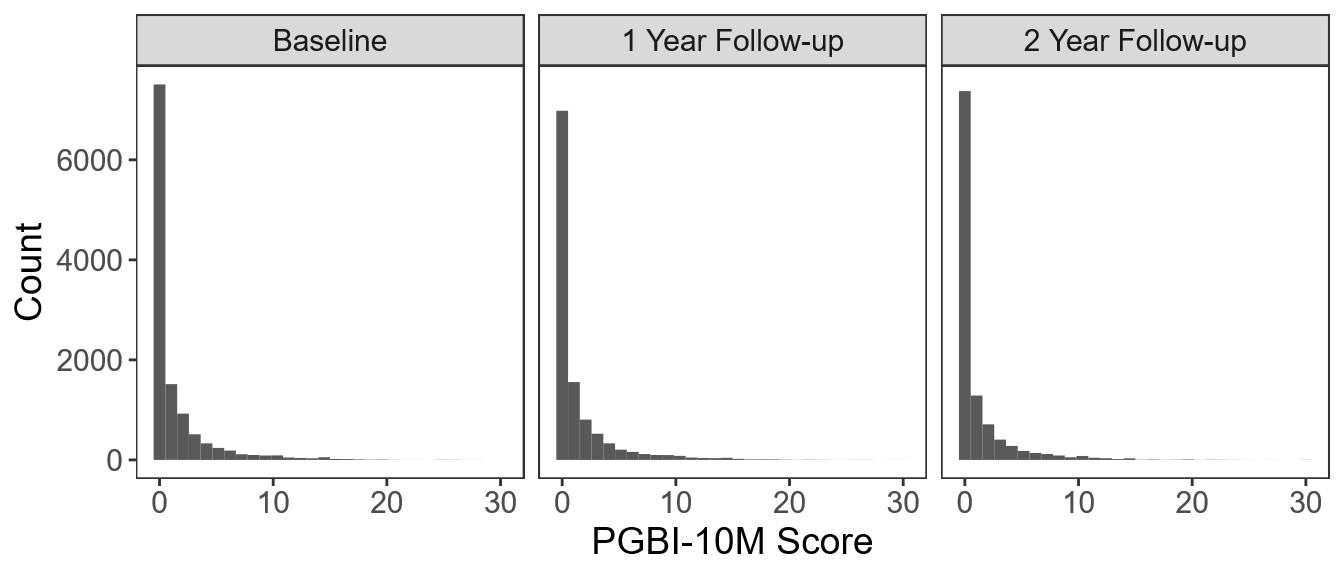

### Supplemental Figure 2. Distributions of mania symptoms for each trajectory at each timepoint

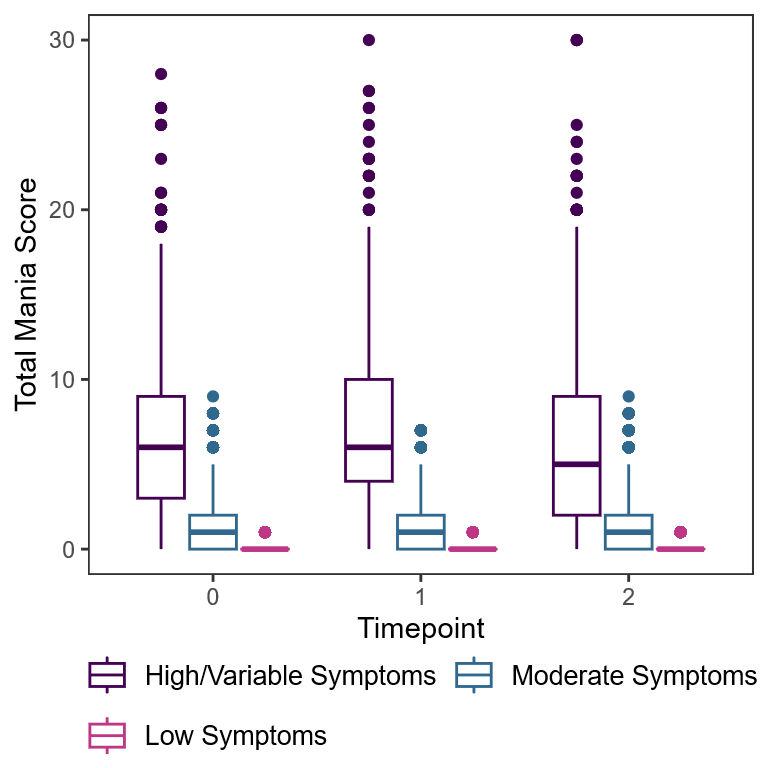

Boxplots indicate median (middle line), interquartile range (box), and outliers (dots) of distributions.

### Supplemental Figure 3. Alternative solutions to growth mixture models of mania symptom trajectories

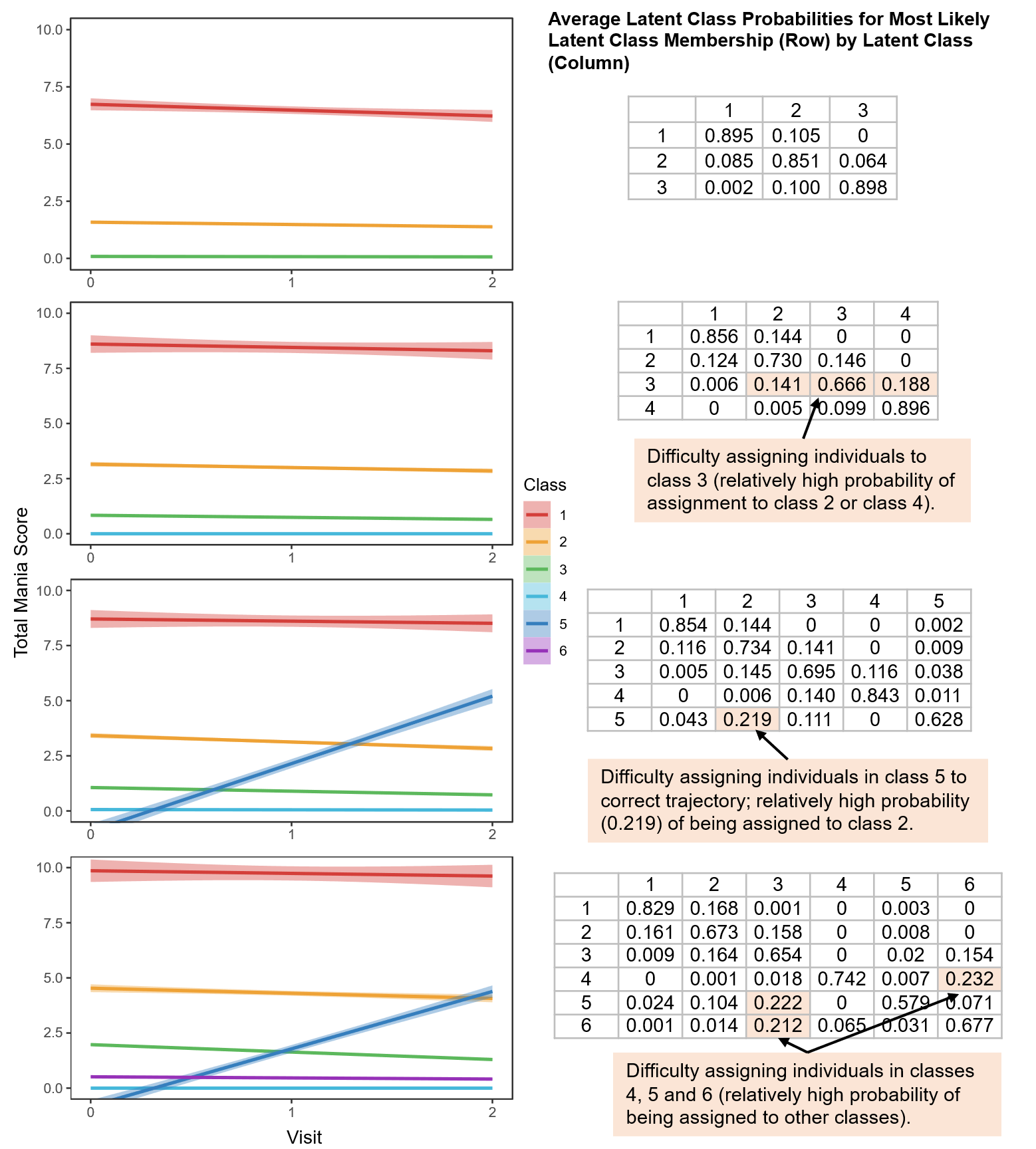
Growth mixture models of mania symptom trajectories comprising 3, 4, 5 and 6 trajectories. Left, plots show mean mania scores for each trajectory class (bolded lines) with 95% confidence intervals (shading). Right, tables show average class probabilities (columns) for individuals assigned to each class (rows). Corresponding fit statistics for each model are presented in Table 2.

### Supplemental Figure 4. Individual symptom scores among individuals in the High/Variable Symptoms trajectory

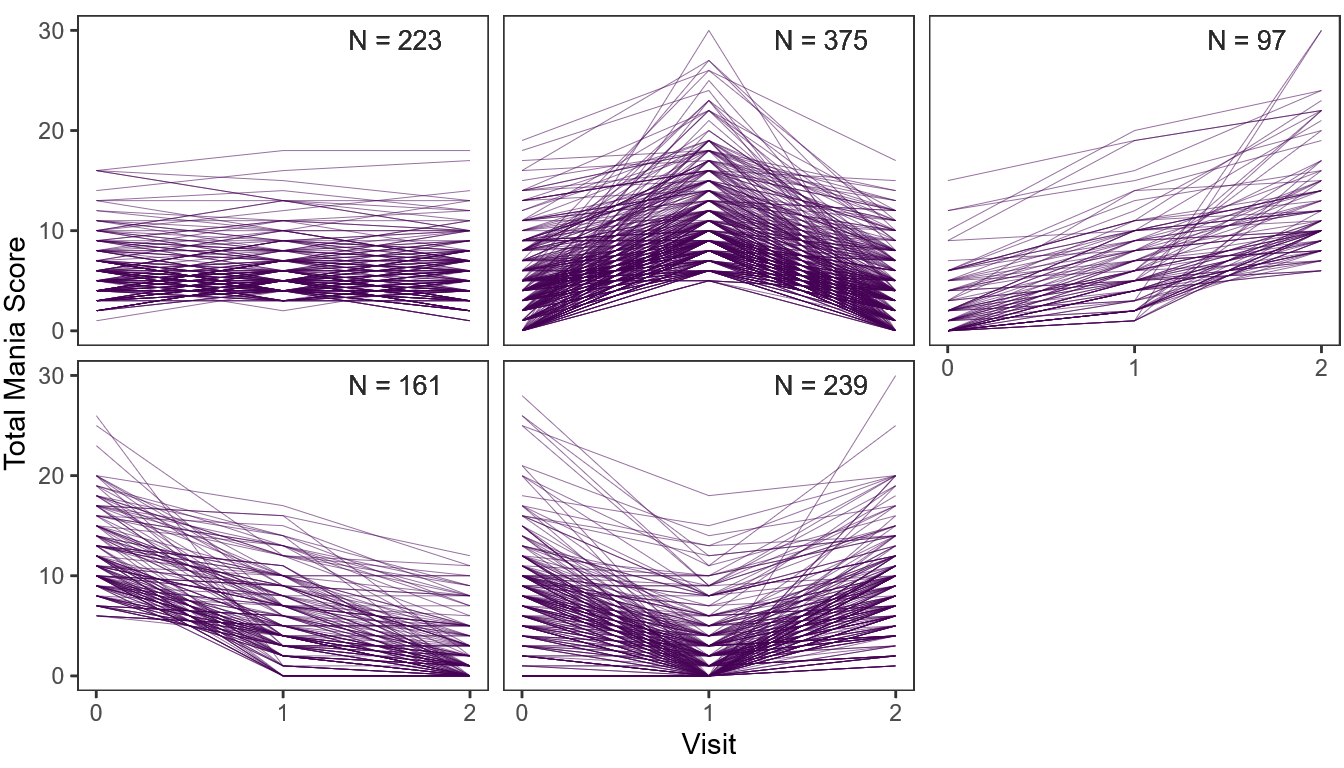

Panels show total mania symptom scores among individuals within the High/Variable Symptoms trajectory (N=1,111). Lines represent individual participants.

### Supplemental Figure 5. Results from joint models testing associations between combinations of risk factors and mania trajectory membership

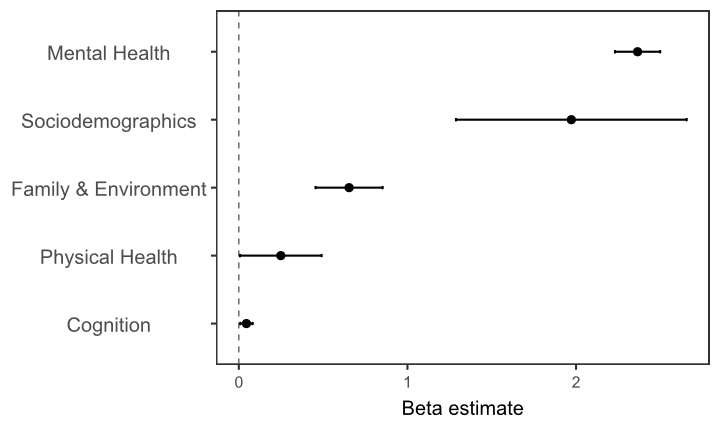

Plot shows beta coefficients from multivariable models testing associations between combinations of risk factors and mania trajectory group membership (represented as a latent variable). Points indicate beta coefficients; bars indicate standard errors. The Mental Health beta coefficient is the sum of coefficients for depression, oppositional defiant disorder, attention deficit hyperactivity disorder, conduct disorder, and anxiety. The Sociodemographic coefficient comprises the summed coefficients for race, ethnicity, socioeconomic status, and sex. The Family & Environment coefficient is the sum of coefficients for first-degree relatives with mania, trauma, maltreatment and family conflict. The Physical Health beta coefficient is the summed coefficients for irritable bowel syndrome, pregnancy complications, obstetric complications, asthma, sleep disturbances, body mass index, cesarean section, premature birth, and low birth weight. The Cognition beta coefficient is the coefficient for verbal learning impairment.

### Supplemental Figure 6. Associations between mania trajectories and sleep disturbance subscales

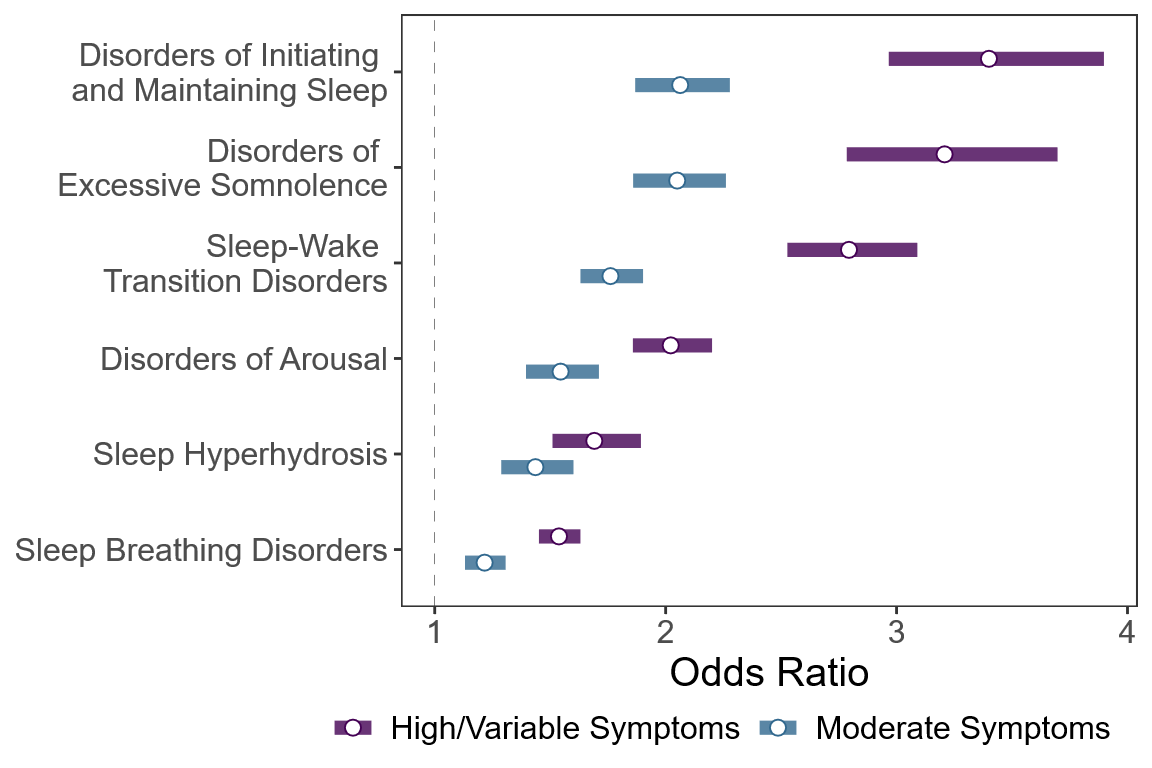

The No Symptoms class serves as the reference group. Circles represent Odds Ratios, bars represent 95% confidence intervals. All models are adjusted for sex, race, ethnicity, and socioeconomic status.
